# Increase in anticholinergic burden in the UK from 1990 to 2015: a UK Biobank study

**DOI:** 10.1101/2020.10.16.20213884

**Authors:** J. Mur, S. R. Cox, R. E. Marioni, G. Muniz-Terrera, T. C. Russ

**Affiliations:** Lothian Birth Cohorts group, Department of Psychology, University of Edinburgh, Edinburgh, UK; Centre for Genomic and Experimental Medicine, Institute of Genetics and Molecular Medicine, University of Edinburgh, Edinburgh, UK; Alzheimer Scotland Dementia Research Centre, University of Edinburgh, Edinburgh, UK; Edinburgh Dementia Prevention, University of Edinburgh, Edinburgh, UK; Division of Psychiatry, Centre for Clinical Brain Science, University of Edinburgh, Edinburgh, UK

## Abstract

**Background:** The use of prescription drugs with anticholinergic properties has been associated with multiple negative health outcomes in older people. Moreover, recent evidence suggests that associated adverse effects may occur even decades after stopping anticholinergic use. Despite the implicated importance of examining longitudinal patterns of anticholinergic prescribing for different age groups, few such data are available.

**Methods:** We performed an age-period-cohort analysis to study trends in anticholinergic burden between the years 1990 and 2015 utilising data from >220,000 UK Biobank participants with linked prescription data from primary care.

**Results:** Anticholinergic burden in the sample increased between three- and nine-fold over 25 years and was significant for both period/cohort- and age-effects across all models. When adjusted for total number of prescriptions, the effect of age reversed. Anticholinergic burden was also associated with various lifestyle- and demographic factors.

**Conclusions:** The increase in anticholinergic prescribing is mostly due to an increase in polypharmacy and is attributable to both ageing of participants, as well as period/cohort-related changes in prescribing practices. There is evidence for deprescribing of anticholinergic medications in older age. Further research is needed to clarify the implications of rising anticholinergic use for public health and to contextualise this rise in light of other relevant prescribing practices.

---

Medicines with anticholinergic properties – antagonists to muscarinic receptors in the nervous system – are commonly prescribed and found among various classes of drugs^1^. Several anticholinergic drugs are listed in the American Geriatrics Society Beers Criteria for Potentially Inappropriate Medication^2^ and the STOPP/START criteria for potentially inappropriate prescribing^3^. Older individuals take more drugs^4^, many of which have anticholinergic effects, and are more sensitive to their side effects^5^. Hence, they are most susceptible to an increased anticholinergic burden.

Anticholinergic burden in older adults is associated with reduced physical and cognitive ability^6,7^, impaired ability to perform activities of daily living^8^, increased risk of dementia^9^ and mortality^10^. The association with dementia has been observed even when the anticholinergic exposure occurred decades prior to diagnosis^9^.

Following the development of the Beers Criteria and other measures^11^, inappropriate prescribing has decreased in the US^12^ and in Europe^13^, though the frequency of anticholinergic prescribing in older adults remains high^14^. Moreover, while anticholinergic prescribing may have decreased in the US^15^, European findings suggest an increase^16–18^. While some studies have found associations between anticholinergic use and demographic factors^15,18,19^, these variables are rarely examined in detail. Moreover, it is not known whether these potential group differences persist over time.

The study of changes over time of prescribing practices with in-depth assessment of age-, period-, and cohort (APC) effects necessitates longitudinal designs. Cross-sectional studies^14^ or repeated cross-sectional studies^16,18^ have explored the extent of anticholinergic use in European countries^16,18^, but the last year of sampling in the UK was in 2010^18^. Moreover, they either lack longitudinal data or rely on participants from a relatively limited geographic area and within a narrow age range. In this paper, we address those limitations by using a large national sample from UK Biobank to characterise longitudinal prescribing patterns of anticholinergic drugs in 1990-2015.

## Methods

### Sample

UK Biobank is a prospective study of >500,000 participants aged 37-73 years recruited UK-wide in 2006-10^20^. Primary care prescription data – including dates, drug codes (BNF/Read v2/CTV3/dm+d) and names – are available for ∼230,000 participants to May 2017 for Scotland, September 2017 for Wales, and August 2017 for England. The data were provided to UK Biobank by region-specific data providers.

### Assignment of anticholinergic burden and drug class

We identified multiple scales^21–30^ from a systematic review^31^; apart from two^27,30^ – all had a four-point (0-3) scoring system of anticholinergic potency (**Table S1**). We derived a meta-scale (**Table S2**) by calculating the mean anticholinergic burden across all 9 original scales that had rated a drug. Thus, scales that scored a drug (even if that score was zero) were included in the computation for that drug, while scales that did not score the drug were not. All prescriptions of medicines with ophthalmic, otic, nasal, or topical routes of administration were assigned an anticholinergic score of zero, as has been done before^26,28,31,32^.

For prescriptions that did not list any drugs, Read-codes – a coded thesaurus of clinical terms used in primary care – were used to supplement them. A series of steps was taken to exclude incomplete data or low number of individuals (**Figures S1, S2**). Drugs were classified based on the Anatomical Therapeutic Chemical (ATC) Classification System (https://www.whocc.no) representing the: (1) anatomical target; (2) therapeutic subgroup; (3) pharmacological subgroup; (4) chemical subgroup; and (5) chemical substance. For example, metformin (5) affects the alimentary tract and metabolism (1), treats diabetes (2), lowers blood-glucose (3), and is a biguanide (4). Not all classes were equally represented in the sample. To allow for comparability of frequency of occurrence, we classified anticholinergic drugs into classes that do not all correspond to the same level in the ATC-hierarchy (see number in the brackets): “drugs for acid disorders” (3), “analgesics” (2), “antidepressants” (3), “antithrombotic drugs” (2), “cardiovascular drugs” (1), “drugs for diabetes” (2), “gastrointestinal drugs” (2), “psycholeptics” (2), “respiratory drugs” (1), and “urological drugs” (3). A class of “other drugs” contained drug categories that individually contributed the least to anticholinergic burden and included anticonvulsants, antibiotics, anti-Parkinsonian drugs, corticosteroids, immunosuppressants, anti-inflammatory drugs, muscle relaxants, and anti-diarrhoeal drugs.

### Statistical approach

To enable longitudinal analyses, the original format of the data was transformed (**Figure S3**). For the analysis of age-period-cohort (APC) effects, we ran three models, excluding one of the APC terms at a time (i.e., its effect was assumed zero). Thus, anticholinergic burden was modelled as a function of either period and cohort (period-cohort model), age and cohort (age-cohort model), or age and period (age-period model). We additionally computed the above models by fitting separate intercepts and slopes: for the period-cohort- and age-cohort models separate intercepts and slopes for each cohort, and for the age-period model separate intercepts and slopes for each period. For analyses of lifestyle- and demographic factors, we fitted Tobit zero-inflated linear models to average monthly anticholinergic burden, adjusting for sex, education, physical activity, social deprivation, region, smoking, BMI, frequency of alcohol consumption, and age at assessment. Outlier observations were removed prior to analysis. For models with random effects, we used the generalized linear mixed models using template model builder (R package glmmTMB^33^); for all other models, we used Tobit regression (R package censReg). Due to relative infrequency of anticholinergic drugs, anticholinergic burden was right-skewed, and models were adjusted for zero inflation. The results were corrected for multiple comparisons using the Bonferroni correction and are reported in unstandardized beta coefficients. Data cleaning and statistical analyses were performed in Python version 3.7.4 and R versions 3.4.1 and 3.6.3. The code used for cleaning and modelling the data is available on GitHub (https://github.com/Logos24/Anticholinergic-trends-UK-Biobank).

Data on sex (male vs. female (ref.)), education (graduate degree vs. no graduate degree (ref.)), the Townsend Index of socioeconomic deprivation^34^ (range: -6.3-11.0, higher values indicate greater deprivation), alcohol consumption (1: daily or almost daily (ref.) vs. 2: three or four times a week vs. 3: once or twice a week vs. 4: one to three times a month vs. 5: only on special occasions vs. 6: never), smoking status (current smoker vs. past smoker vs. never smoker (ref.)), BMI, and physical activity (strenuous vs. moderate vs. mild (ref.))^35^ were ascertained during or immediately prior to the participants’ recruitment to UK Biobank. Region (Scotland vs. Wales vs. England(ref.)) was derived by combining data providers so that all prescriptions issued in England, Scotland, and Wales, respectively, were classified under the same class.

Each model was run in three iterations: basic models were unadjusted; basic-adjusted models included sex, data provider, education, and socioeconomic deprivation. Data providers were specific to each prescription and available longitudinally. Sex, education, and deprivation were assumed constant within individuals: over 90% of UK Biobank participants reported the same educational attainment at reassessment; within-person stability of deprivation has been reported previously^36^. Fully-adjusted models were additionally controlled for smoking, alcohol consumption frequency, BMI, and physical activity. While these covariates were available only cross-sectionally, they are important in health and disease.

## Results

The 220,867 participants were born 1938-1969 (**Suppl. Figure 4**). Individuals were continuously recruited, but the demographic structure of the sample (**Table 1**) remained relatively stable over time. However, it is unclear how demographic variables changed within individuals over time.

**Table 1:**
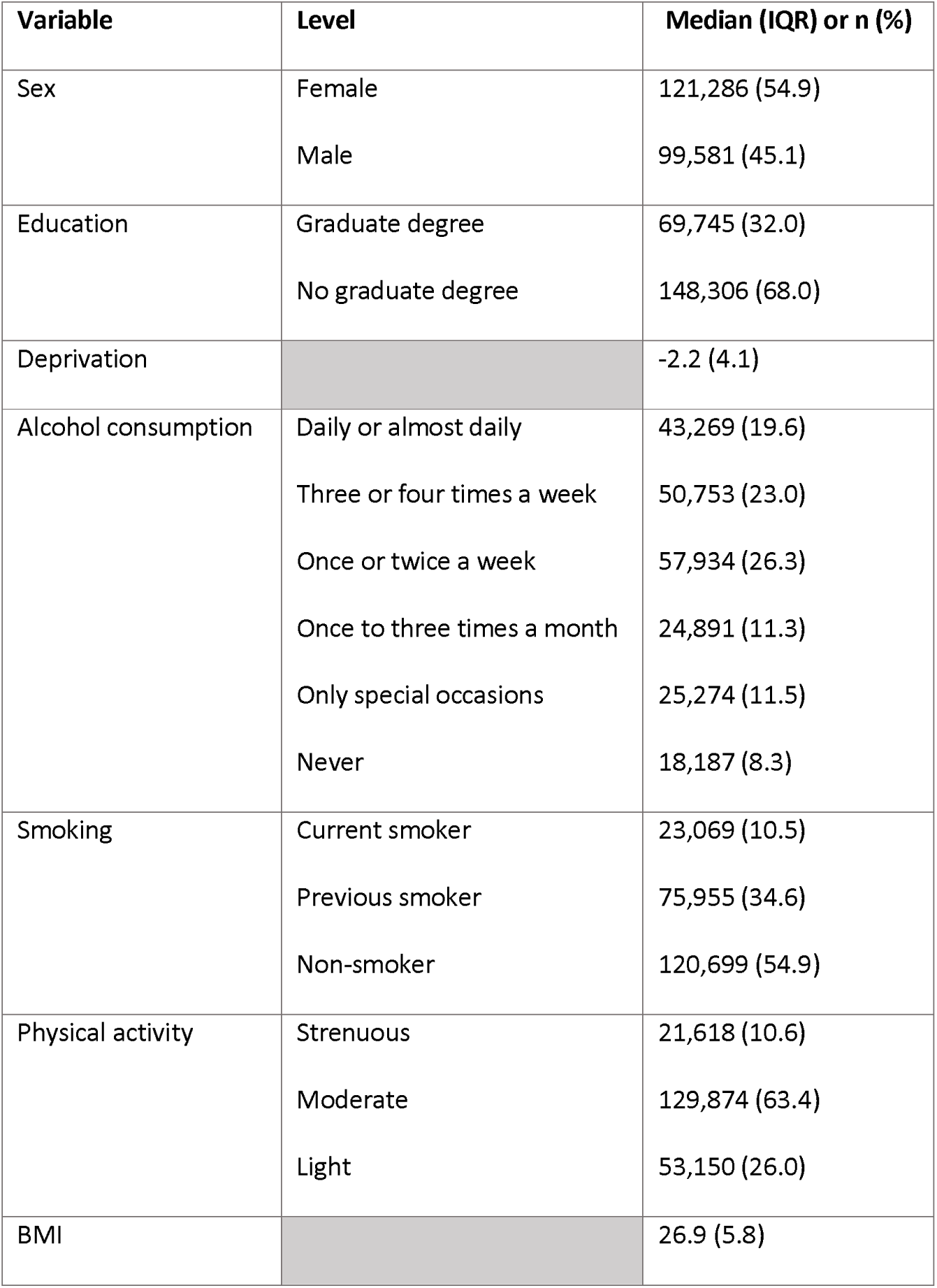
Demographic characteristics of the sample at the time of recruitment to UK Biobank.

| Variable | Level | Median (IQR) or n (%) |
| --- | --- | --- |
| Sex | Female | 121,286 (54.9) |
|  | Male | 99,581 (45.1) |
| Education | Graduate degree | 69,745 (32.0) |
|  | No graduate degree | 148,306 (68.0) |
| Deprivation |  | -2.2 (4.1) |
| Alcohol consumption | Daily or almost daily | 43,269 (19.6) |
|  | Three or four times a week | 50,753 (23.0) |
|  | Once or twice a week | 57,934 (26.3) |
|  | Once to three times a month | 24,891 (11.3) |
|  | Only special occasions | 25,274 (11.5) |
|  | Never | 18,187 (8.3) |
| Smoking | Current smoker | 23,069 (10.5) |
|  | Previous smoker | 75,955 (34.6) |
|  | Non-smoker | 120,699 (54.9) |
| Physical activity | Strenuous | 21,618 (10.6) |
|  | Moderate | 129,874 (63.4) |
|  | Light | 53,150 (26.0) |
| BMI |  | 26.9 (5.8) |

### Anticholinergic prescribing

Of 248 drugs on the meta-scale, 201 (81.0%) were found in the sample and constituted 25.0% of all prescriptions. A total of 199,652 participants (90.4%) were prescribed at least one anticholinergic drug and 28,525 (13.2%) participants were prescribed anticholinergic drugs every year during the prescribing period. Among previously published scales, anticholinergic prescriptions constituted 2.5%-23.1% of all prescriptions **(Table 2)** and anticholinergic burden according to each scale exhibited an increasing trend over time **(Figure 1A**). Depending on the scale used, anticholinergic burden increased between three- and nine-fold from 1990 to 2015. Most anticholinergic prescriptions were for antidepressants, which accounted for 32.5% of the total anticholinergic burden. **(Table 3, Figure 1B**). The anticholinergic burden for each drug class increased with time **(Figure 1C)**.

**Table 2:**
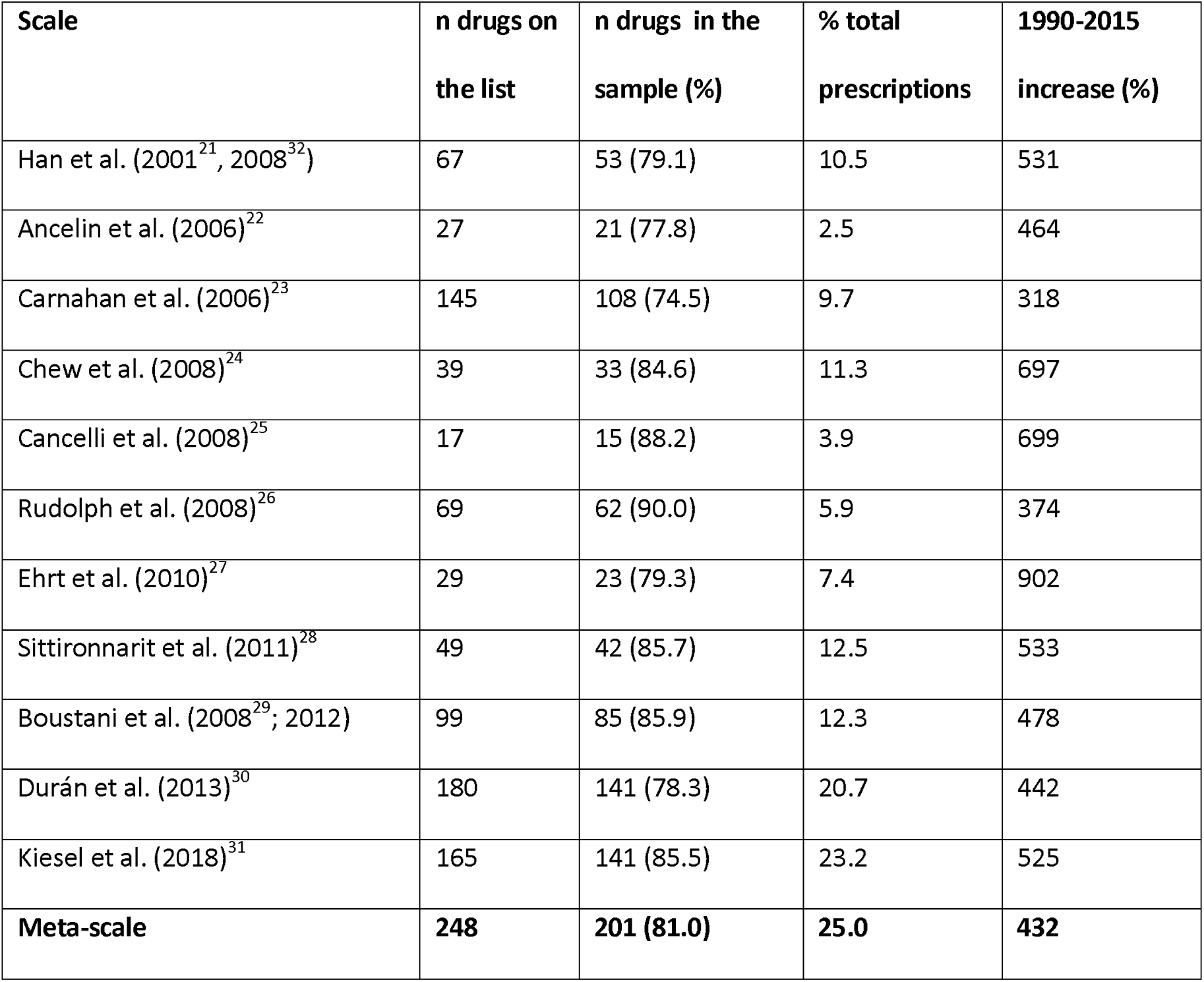
Comparison of the nu mbers of drugs on each anticholinergic scale, the numbers of drugs on each scale that were prescribed in our sample, the percentages of all prescriptions in the sample that the drugs on the scales constituted, and the increase in the mean yearly anticholinergic burden from the year 1990 to the year 2015.

| Scale | n drugs on the list | n drugs in the sample (%) | % total prescriptions | 1990-2015 increase (%) |
| --- | --- | --- | --- | --- |
| Han et al. (2001<sup>21</sup>, 2008<sup>32</sup>) | 67 | 53 (79.1) | 10.5 | 531 |
| Ancelin et al. (2006)<sup>22</sup> | 27 | 21 (77.8) | 2.5 | 464 |
| Carnahan et al. (2006)<sup>23</sup> | 145 | 108 (74.5) | 9.7 | 318 |
| Chew et al. (2008)<sup>24</sup> | 39 | 33 (84.6) | 11.3 | 697 |
| Cancelli et al. (2008)<sup>25</sup> | 17 | 15 (88.2) | 3.9 | 699 |
| Rudolph et al. (2008)<sup>26</sup> | 69 | 62 (90.0) | 5.9 | 374 |
| Ehrt et al. (2010)<sup>27</sup> | 29 | 23 (79.3) | 7.4 | 902 |
| Sittironnarit et al. (2011)<sup>28</sup> | 49 | 42 (85.7) | 12.5 | 533 |
| Boustani et al. (2008<sup>29</sup>; 2012) | 99 | 85 (85.9) | 12.3 | 478 |
| Durán et al. (2013)<sup>30</sup> | 180 | 141 (78.3) | 20.7 | 442 |
| Kiesel et al. (2018)<sup>31</sup> | 165 | 141 (85.5) | 23.2 | 525 |
| **Meta-scale** | **248** | **201 (81.0)** | **25.0** | **432** |

**Table 3:** Comparison of the num bers of anticholinergic drugs from different drug classes and their contributions to the total anticholinergic burden.

| **Drug class** | **n** | **%** | **Number of drugs in class** | **Mean anticholinergic burden per drug** | **% total anticholinergic burden** |
| --- | --- | --- | --- | --- | --- |
| Acid disorders | 1,464,542 | 10.3 | 5 | 0.50 | 5.8 |
| Analgesics | 1,426,703 | 10.1 | 12 | 1.48 | 16.5 |
| Antidepressants | 2,678,379 | 18.9 | 27 | 1.54 | 32.4 |
| Antithrombotics | 546,311 | 3.8 | 2 | 0.41 | 1.8 |
| Cardiovascular | 2,006,594 | 14.1 | 16 | 0.53 | 8.4 |
| Diabetes | 785,940 | 5.5 | 1 | 0.38 | 2.3 |
| Gastrointestinal | 220,269 | 1.6 | 9 | 1.18 | 2.0 |
| Psycholeptic | 690,894 | 4.9 | 35 | 1.16 | 6.3 |
| Respiratory | 1,499,455 | 10.6 | 33 | 0.88 | 10.3 |
| Urological | 330,314 | 2.3 | 8 | 2.41 | 6.2 |
| Other | 2,545,293 | 17.9 | 57 | 0.40 | 8.0 |

**Figure 1:**
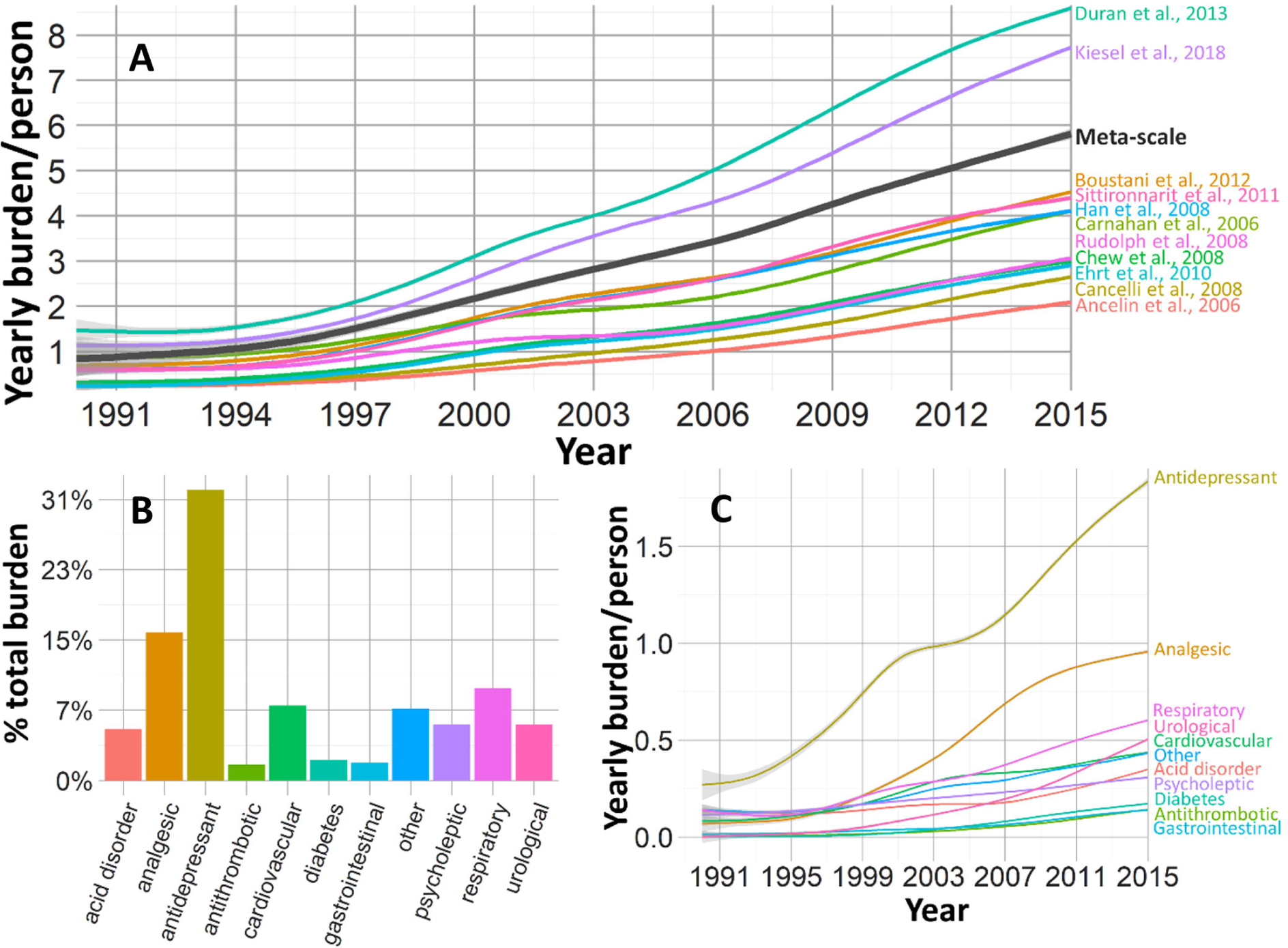
Anticholinergic burden over time based on different anticholinergic scales **(A)**, percentage of anticholinergic burden in the sample due to each drug class **(B)**, and the change in anticholinergic burden over time due to each drug class **(C)**. The plots in **A** and **B** were generated using generalised additive model smoothing.

### Age-Period-Cohort-analysis

In the basic period-cohort model, anticholinergic burden was positively associated with period (beta=0.0095, SE=9.6×10^−6^, p<2.0×10^−16^) and negatively associated with cohort (beta=-0.065, SE=8.9×10^−5^, p<2.0×10^−16^). In the basic age-cohort model, anticholinergic burden was positively associated with age (beta=0.11, SE=1.1×10^−4^, p<2.0×10^−16^) and with cohort (beta=0.049, SE=1.4×10^−4^, p<2.0×10^−16^). In the basic age-period model, anticholinergic burden was positively associated with age (beta=0.065, SE=8.9×10^−5^, p<2.0×10^−16^) and with period (beta=0.0040, SE=1.1×10^−5^, p<2.0×10^−16^). The same trends were observed in the basic-adjusted- and fully-adjusted models **(Suppl. Table 3**). These results indicate that greater anticholinergic burden relates to both ageing and later chronological time (period) and/or earlier-cohort. That is, in a given period, older individuals experience a higher anticholinergic burden than younger individuals in the same period. Moreover, in recent periods, individuals will experience a higher anticholinergic burden than individuals of the same age did in the past. For example, the average yearly anticholinergic burden of a 50-year old was 2.32 in 2000, 2.92 in 2007, and 3.67 in 2015, while the average yearly anticholinergic burden of a 60-year-old was 3.06 in 2000, 3.94 in 2007, and 5.12 in 2015.

The results were also significant in the mixed-effect models. In the basic period-cohort model, period was positively associated with anticholinergic burden (beta=0.13, SE=0.0012, p<2.0×10^−16^) and earlier-born cohorts exhibited steeper slopes than later-born cohorts (correlation between slope and cohort: n=32, r=-0.97, 95% CI=-0.98—0.95, p<2.0×10^−16^; **Figure 2A**). In the age-cohort model, age was positively associated with anticholinergic burden (beta=0.13, SE=0.0012, p<2.0×10^−16^; **Figure 2B)** and earlier-born cohorts exhibited steeper slopes than later-born cohorts (n=32, correlation between slope and cohort: r=-0.97, 95% CI=-0.98--0.93, p<2.0×10^−16^). In the age-period model, age was positively associated with anticholinergic burden (beta=0.13, SE=0.0012, p<2.0×10^−16^; **Figure 2C)** and later periods exhibited steeper slopes than earlier periods (n=25, correlation between slope and period: r=0.95, 95% CI=0.89-0.98, p=1.3×10^−13^). The trends persisted when the outcome was the number of prescribed anticholinergic drugs **(Suppl. Table 4**). The proportion of drugs with different anticholinergic potencies remained stable over time (**Suppl. Figure 5**). Thus, the increase in anticholinergic burden was likely due to a general increase in anticholinergic prescribing, rather than a relative increase in the prescribing of stronger anticholinergic drugs.

**Figure 2:**
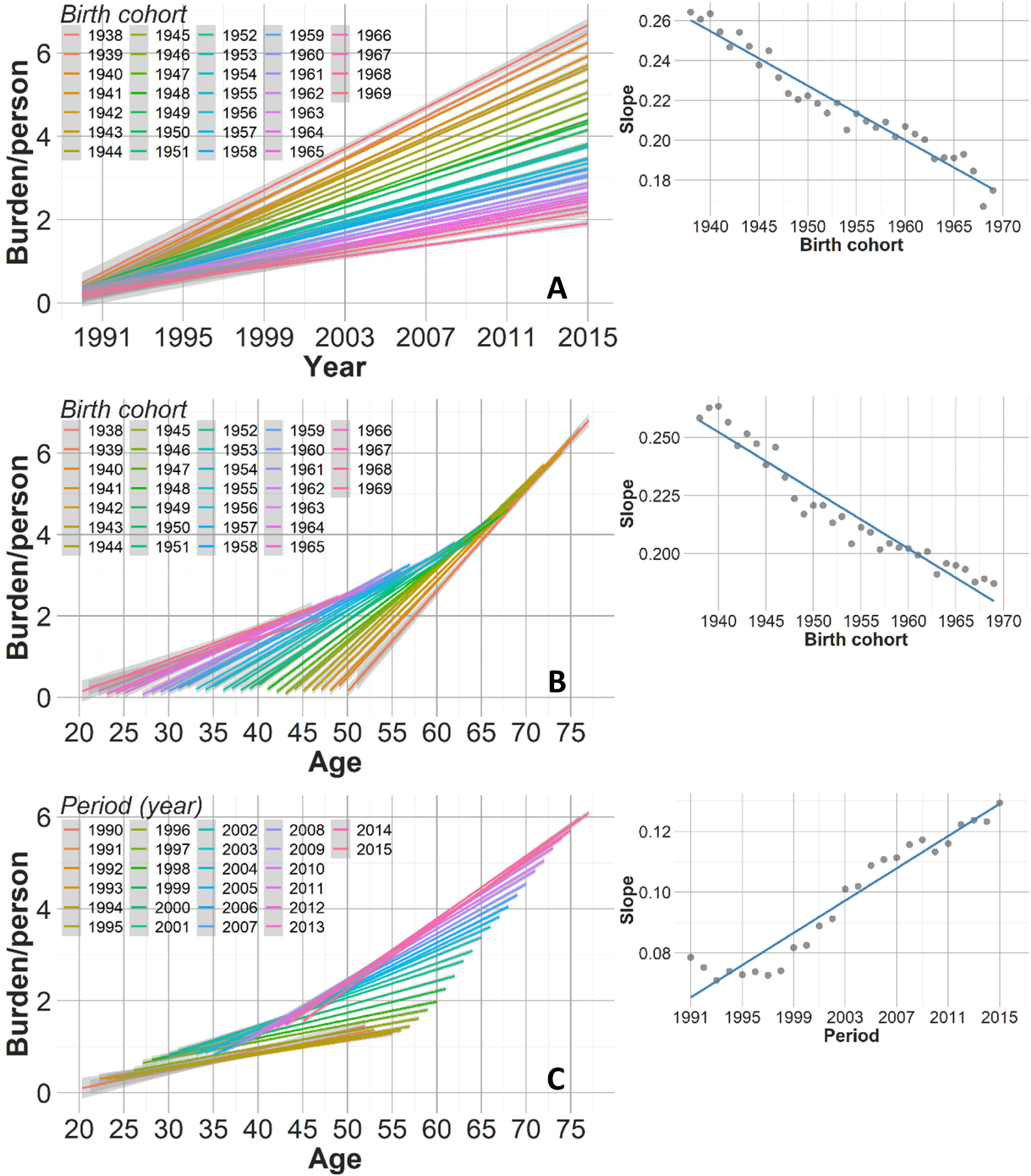
APC-analysis with basic mixed models with random intercepts and slopes **(left)** and associations between slopes and different levels of predictors **(right). A** depicts the period-cohort model with cohort as a random effect, **B** depicts the age-cohort model with cohort as a random effect, and **C** depicts the age- period model with period as the random effect.

When the change in anticholinergic burden was plotted for each drug class separately **(Figure S6)**, the same pattern was observed for all drug classes except for drugs for acid disorders and cardiovascular drugs. For the former, an increase in anticholinergic burden over time was observed, but was similar across periods and cohorts, suggesting an effect of age, but without a prominent cohort/period effect. For cardiovascular drugs, we observed an increase in anticholinergic burden over time and a higher anticholinergic burden in earlier cohorts and later periods, suggesting a positive effect of age, but a negative period-effect.

When the basic models were adjusted by the addition of the total number of prescribed drugs (**Table S5**), the effect of birth cohort was reversed in the period-cohort model and the effect of age was reversed in the age-period model. In the former, anticholinergic burden was positively associated with period and with birth cohort; in the latter, anticholinergic burden was negatively associated with age and positively associated with period. These results indicate the retention of the period/cohort effect, but a reversal of the effect of age when adjusted for the number of prescribed drugs. Thus, whereas overall anticholinergic burden has increased over time, and more so among older adults, anticholinergic drugs in the latter group comprise a relatively lower proportion of overall prescriptions when compared to younger individuals **(Suppl. Figure 7)**.

### Anticholinergic burden and demographic factors

Higher anticholinergic burden was associated with female sex, lower educational attainment, greater deprivation, and higher BMI, less frequent alcohol consumption, lower physical activity, and was greater in Scotland and Wales than in England **(Table 5)**.

**Table 5:**
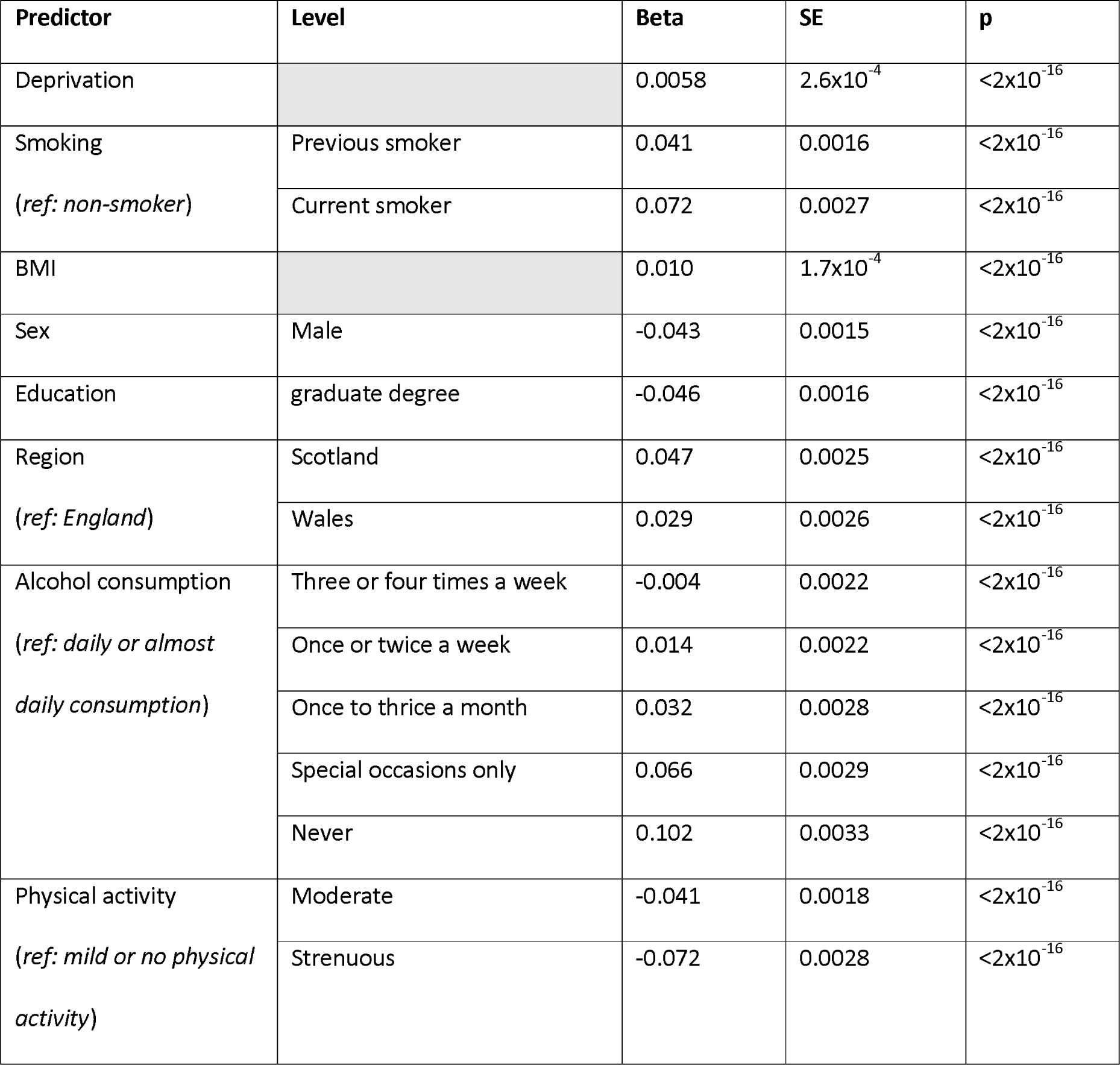
Results of the model predicting monthly anticholinergic burden as a function of deprivation, smoking, BMI, sex, education, region, alcohol consumption, physical activity, and age.

| Predictor | Level | Beta | SE | p |
| --- | --- | --- | --- | --- |
| Deprivation | | 0.0058 | $2.6 \times 10^{-4}$ | $< 2 \times 10^{-16}$ |
| Smoking<br>(*ref: non-smoker*) | Previous smoker | 0.041 | 0.0016 | $< 2 \times 10^{-16}$ |
| | Current smoker | 0.072 | 0.0027 | $< 2 \times 10^{-16}$ |
| BMI | | 0.010 | $1.7 \times 10^{-4}$ | $< 2 \times 10^{-16}$ |
| Sex | Male | -0.043 | 0.0015 | $< 2 \times 10^{-16}$ |
| Education | graduate degree | -0.046 | 0.0016 | $< 2 \times 10^{-16}$ |
| Region<br>(*ref: England*) | Scotland | 0.047 | 0.0025 | $< 2 \times 10^{-16}$ |
| | Wales | 0.029 | 0.0026 | $< 2 \times 10^{-16}$ |
| Alcohol consumption<br>(*ref: daily or almost daily consumption*) | Three or four times a week | -0.004 | 0.0022 | $< 2 \times 10^{-16}$ |
| | Once or twice a week | 0.014 | 0.0022 | $< 2 \times 10^{-16}$ |
| | Once to thrice a month | 0.032 | 0.0028 | $< 2 \times 10^{-16}$ |
| | Special occasions only | 0.066 | 0.0029 | $< 2 \times 10^{-16}$ |
| | Never | 0.102 | 0.0033 | $< 2 \times 10^{-16}$ |
| Physical activity<br>(*ref: mild or no physical activity*) | Moderate | -0.041 | 0.0018 | $< 2 \times 10^{-16}$ |
| | Strenuous | -0.072 | 0.0028 | $< 2 \times 10^{-16}$ |

Examining each drug class separately, most effects remained **(Suppl. Table 6)**. However, anticholinergic burden due to antithrombotic drugs, cardiovascular drugs, and drugs for diabetes was higher in males than in females. Moreover, regional differences in anticholinergic burden strongly depended on drug class. For region, sex, education, and deprivation, we plotted anticholinergic burden as a function of period for different levels of predictor variables **(Suppl. Figure 8**). For this purpose, deprivation was transformed into a binary categorical variable, with the median (∼-2.2) across all participants as the boundary for deprivation. Visual inspection confirmed the effects of the predictors on anticholinergic burden. Furthermore, the disparities in anticholinergic burden between the levels of categorical predictors seemed to increase over time.

## Discussion

Anticholinergic burden in the UK is increasing and older individuals continue to have the highest anticholinergic burden. Age-related increases in anticholinergic burden can be explained by polypharmacy in older adults. Indeed, when accounting for polypharmacy and period, anticholinergic burden decreases with age, possibly demonstrating proportionate deprescribing of anticholinergic drugs in older age. We also find associations between higher anticholinergic burden and various demographic- and lifestyle factors, including female sex, less education, and greater socioeconomic deprivation.

### Anticholinergic burden over time

Anticholinergic burden increased in all APC models. Throughout time periods and across birth cohorts, ageing was associated with greater anticholinergic burden. Moreover, across age groups and birth cohorts, anticholinergic burden has increased in recent years. Finally, at a given age, later-born cohorts experienced a greater anticholinergic burden than earlier-born cohorts, while in a given period, later born cohorts experienced a smaller anticholinergic burden than earlier-born cohorts.

Because of the collinearity of age, period, and cohort (age=period-cohort), they cannot all be included in a regression analysis, as holding two terms constant keeps the third term constant as well^37^. Some argue that the APC problem cannot be completely resolved^38^ and that results from APC-based models should be based on well-founded and clearly communicated assumptions. It is that approach that we adopted in the present paper and based on current knowledge on polypharmacy and anticholinergic burden, some conclusions can be reached. First, due to increased multimorbidity and polypharmacy in older individuals^4^, age probably contributed to the trend. When intercept and slope were modelled separately in mixed models with random effects, cohort was negatively associated with the slope, suggesting not only a greater anticholinergic burden, but also a more rapid accumulation of burden in older individuals. Second, as previously reported^4^, individuals are now being prescribed more drugs than in the past. The increase in anticholinergic burden could be caused by a new generation of patients who either demand more or who are diagnosed with more maladies. Alternatively, the increase could be related to changes in prescribing practices due to societal changes or changes in medical training. Regardless of the underlying causes, people in the UK are being increasingly prescribed anticholinergic drugs.

The increases in anticholinergic burden could be related to polypharmacy and not an increase in specifically anticholinergic prescribing. Indeed, when the models were adjusted for the number of prescriptions, earlier-born individuals exhibited a lower anticholinergic burden across periods and across age groups than those born later. Moreover, across age groups, anticholinergic burden was higher in later periods than in earlier periods. While correcting for polypharmacy had no effect on the trend of the age-cohort model, it changed the direction of birth cohort and age in the period-cohort model and the age-period model, respectively. Later-born individuals exhibited a higher anticholinergic burden, and this burden was positively associated with period, but negatively associated with age. While this indicates that medical practitioners have been mitigating the increase in polypharmacy by deprescribing anticholinergic drugs in older people, this group nevertheless experienced the highest burden. Furthermore, older people experienced a greater anticholinergic burden in 2015 than at any point in the preceding 25 years.

### Demographic- and lifestyle factors

Anticholinergic use has been linked with some demographic- and lifestyle factors^15,18,19^. In our study, female sex, lower education, higher socioeconomic deprivation, higher BMI, lower frequency of alcohol consumption, lower physical activity, and being prescribed in Scotland or Wales (compared to England) were associated with a higher anticholinergic burden. Certain groups do require a greater number of medications but medical professionals may prescribe more to certain groups, independent of underlying medical conditions.

Interestingly, greater alcohol consumption was associated with decreased anticholinergic burden. Individuals who take many medications may reduce their alcohol consumption to reduce the risk of drug interactions or to reduce the impact of existing disease.

### Strengths and limitations

The present study used a very large, well-characterised sample and utilised primary care electronic prescription data over a wide period. However, we recognise several limitations. First, while visual inspection of anticholinergic burden across different scales did reveal a common upward trend, our newly computed meta-scale was not previously validated and likely overestimates anticholinergic burden. Second, we did not include longitudinal data on over-the-counter drugs and dietary supplements. Third, while our assumption that topical, ophthalmic, otic, and nasal drugs do not have anticholinergic effects is common in the literature^26,28,31,32^, we are not aware of conclusive evidence to support it. Fourth, estimates of prevalence and statistical inferences are dependent on the underlying sample and UK Biobank is not representative of the UK population. On average, participants in the study are less likely to be obese, to smoke, have fewer health conditions, and live in socioeconomically less deprived areas^39^. Thus, differences in anticholinergic burden and period-dependent disparities are possibly greater in real populations. Fifth, our analysis of the effects of demographic and lifestyle factors on anticholinergic burden assessed the correlation between the average value of a metric that changes with time (anticholinergic burden) and cross-sectional data (e.g. BMI) which was ascertained towards the end of the period in question, when participants were of different ages. Thus, our results cannot clarify the exact nature of the temporal relationship. Finally, we did not have data on the oldest old, who represent the group most at risk by anticholinergic effects.

### Conclusion and future directions

Prescribing drugs involves balancing their medicinal value with potential harms. Moreover, exhaustive longitudinal studies are required to fully determine all their effects. However, based on the current evidence, anticholinergic drugs ought to be prescribed sparingly, and the use of alternatives strongly considered. An understanding of temporal prescribing trends in a population may help to guide prescribing and stimulate further research. Our work represents an overview and future studies should describe prescribing trends and their relationship to age groups, and demographic- and lifestyle characteristics in greater detail. There is also evidence of differences between drug classes in the association between anticholinergic burden and health outcomes^9^. Identifying distinct anticholinergic trends for individual drug classes for different groups could help to further improve prescribing guidelines. Additionally, future work should attempt to identify the causes for the increase in anticholinergic prescribing, and more precisely quantify the potential implications for important life outcomes including brain- and cognitive health, and dementia. Finally, decreases in potentially inappropriate prescribing have been reported even when the same population experienced increases in polypharmacy and in anticholinergic use^17^. Thus, increases in anticholinergic burden should not be considered in isolation, but in the context of other prescribing practices.

## Data Availability

All raw data is available via UK Biobank.

https://github.com/Logos24/Anticholinergic-trends-UK-Biobank

## Acknowledgements

REM is supported by Alzheimer’s Research UK major project grant ARUK-PG2017B-10. JM is supported by funding from the Wellcome Trust 4-year PhD in Translational Neuroscience—training the next generation of basic neuroscientists to embrace clinical research [108890/Z/15/Z]. JM and TCR are members of the Alzheimer Scotland Dementia Research Centre funded by Alzheimer Scotland. TCR is employed by NHS Lothian and the Scottish Government. SRC is supported by Age UK (Disconnected Mind project), the UK Medical Research Council [MR/R024065/1] and a National Institutes of Health (NIH) research grant R01AG054628. The authors thank all participants of the UK Biobank for providing data for the study, Dr Michelle Luciano (Department of Psychology, University of Edinburgh) for managing UK Biobank data application 10279, and Dr Andrew Bell (Sheffield Methods Institute, University of Sheffield) for input on data analysis. The Research Ethics Committee (REC) granted ethical approval for the study—reference 11/NW/0382—and the current analysis was conducted under data application 10279.

## Notes

### Competing Interest Statement

The authors have declared no competing interest.

### Author Declarations

Research Ethics Committee (REC) (reference 11/NW/0382).

