## Supplemental material for "Increase in anticholinergic burden in the UK from 1990 to 2015: a UK Biobank study"

**Table S1**: Anticholinergics scales identified in the present study. We considered anticholinergic scales that were available as complete lists of drugs, scored each drug for its anticholinergic potency, and did not utilize dosage. Grey shading indicates that the scale was not considered for further analysis. For two scales^1,2^, updated versions were used (Aging Brain Care, 2012; Carnahan, 2014, personal communication on 21.10.2019). One scale^3^ was modified to include newer drugs from the UK market as has been done before^4^. Most scales use a four-point (0-3) scoring system of anticholinergic potency, where 0 indicates no anticholinergic effect and 3 indicates a strong anticholinergic effect. One study^5^ used a two-point system, where 2 and 1 indicated strong and weak anticholinergic scores, respectively. Some drugs from this scale were categorised as “drugs with improbable or no anticholinergic action”. For our analyses, the drugs in the latter category were scored with 1, while the scores of other drugs on the scale were increased by one point. Another study^6^ used a five-point, 0-4 scale, which was changed to a 0-3 scale as has been done before^5,7^.

| **Surname of first author** | **Year of publication** | **Reason for exclusion** |
| --- | --- | --- |
| Summers^8^ | 1978 | Outdated (based on the date of publication and on new scales developed on its basis). |
| Han^9,10^ | 2001 |  |
| Aizenberg^11^ | 2002 | Publicly unavailable and no response from lead author to two email requests within a year. |
| Minzenberg^12^ | 2004 | Based on a reference compound. |
| Ancelin^13^ | 2006 |  |
| Carnahan^1^ | 2006 |  |
| Hilmer^14^ | 2007 | Required information on drug dosage. |
| Chew^15^ | 2008 |  |
| Cancelli^16^ | 2008 |  |
| Rudolph^3^ | 2008 |  |
| Ehrt^6^ | 2010 |  |
| Sittironnarit^17^ | 2011 |  |
| Boustani^2^ | 2008 |  |
| Whalley^18^ | 2012 | Unavailable in full. |
| Durán^5^ | 2013 |  |
| Dauphinot^19^ | 2014 | Required information on drug dosage. |
| Klamer^20^ | 2017 | Required information on drug dosage. |
| Kiesel^7^ | 2018 |  |

**Table S2**: The meta-scale, with generic drug names for all anticholinergic drugs (i.e., drugs with an anticholinergic score greater than 0) listed in the first column and their respective anticholinergic scores in the second column.

| **Drug name** | **Score** |
| --- | --- |
| acepromazine | 3 |
| aceprometazine | 3 |
| aclidinium bromide | 1 |
| alimemazine | 1.5 |
| alprazolam | 1 |
| alverine | 1.5 |
| amantadine | 1.25 |
| aminophylline | 1 |
| amitriptyline | 3 |
| amoxapine | 3 |
| amoxicillin | 0.125 |
| ampicillin | 0.5 |
| aripiprazole | 0.25 |
| asenapine | 1 |
| atenolol | 0.333333 |
| atropine | 3 |
| azatadine | 3 |
| azathioprine | 1 |
| baclofen | 1 |
| barberry | 1 |
| belladonna | 2.75 |
| benazepril | 0.5 |
| benztropine | 3 |
| betaxolol | 0.333333 |
| biperiden | 3 |
| bisacodyl | 0.25 |
| bromocriptine | 0.666667 |
| brompheniramine | 2 |
| buclizine | 3 |
| bupropion | 0.333333 |
| captopril | 0.4 |
| carbamazepine | 0.5 |
| carbidopa | 0.333333 |
| carbidopa/levodopa | 0.5 |
| carbinoxamine | 3 |
| carisoprodol | 1 |
| cefalexin | 0.125 |
| cefamandole | 1 |
| cefoxitin | 1 |
| celecoxib | 0.5 |
| cephalotin | 1 |
| cetirizine | 1.166667 |
| chlordiazepoxide | 1 |
| chloroquine | 1 |
| chlorphenamine | 3 |
| chlorpromazine | 2.833333 |
| chlorprothixene | 3 |
| chlortalidone | 0.5 |
| ciclosporin | 1 |
| cimetidine | 1.666667 |
| citalopram | 1 |
| clemastine | 3 |
| clidinium | 2 |
| clindamycin | 0.5 |
| clomipramine | 2.8 |
| clonazepam | 0.666667 |
| clorazepate | 1.666667 |
| clotiapine | 3 |
| clozapine | 2.833333 |
| codeine | 0.5 |
| colchicine | 1 |
| corticosterone | 1 |
| cortisone | 0.5 |
| cyclizine | 3 |
| cyclobenzaprine | 1.75 |
| cycloserine | 1 |
| cyproheptadine | 2.2 |
| darifenacin | 2.666667 |
| desipramine | 2.5 |
| desloratadine | 0.5 |
| dexamethasone | 0.333333 |
| dexbrompheniramine | 3 |
| dexchlorpheniramine | 3 |
| dextromethorphan | 0.5 |
| diazepam | 0.75 |
| dicycloverine | 2.4 |
| digitoxin | 1 |
| digoxin | 1.0625 |
| dimenhydrinate | 3 |
| dimetindene | 1 |
| diphenhydramine | 2.8 |
| diphenoxylate | 0.166667 |
| diphenoxylate/atropine | 3 |
| dipyridamole | 0.25 |
| disopyramide | 0.75 |
| domperidone | 0.5 |
| donepezil | 0.1 |
| dosulepin | 2.333333 |
| doxepine | 2.857143 |
| doxylamine | 2 |
| duloxetine | 0.166667 |
| emepronium | 3 |
| entacapone | 0.5 |
| ephedrine | 1 |
| ergotamine | 1 |
| escitalopram | 0.75 |
| estazolam | 1 |
| etoricoxib | 1 |
| famotidine | 0.25 |
| fentanyl | 0.5 |
| fesoterodine | 2.666667 |
| fexofenadine | 1 |
| flavoxate | 2.666667 |
| flunitrazepame | 1 |
| flunizepam | 1 |
| fluoxetine | 1 |
| fluphenazine | 2 |
| flurazepam | 1 |
| fluvoxamine | 1 |
| furosemide | 0.9375 |
| gentamicin | 1 |
| glycopyrronium | 3 |
| guaifenesin | 0.5 |
| haloperidol | 0.857143 |
| homatropine | 3 |
| hydralazine | 0.5 |
| hydrocodone | 0.833333 |
| hydrocortisone | 0.5 |
| hydroxyzine | 2.4 |
| hyoscine butylbromide | 3 |
| hyoscine hydrobromide | 3 |
| hyoscyamine | 3 |
| iloperidone | 1 |
| imipramine | 3 |
| ipratropium | 1.25 |
| isosorbide | 1 |
| ketamine | 3 |
| ketorolac | 0.5 |
| ketotifen | 1 |
| lansoprazole | 0.3 |
| levocetirizine | 0.5 |
| levofloxacin | 0.166667 |
| levomepromazine | 2.2 |
| lithium | 1 |
| lofepramine | 1 |
| loperamide | 0.833333 |
| loratadine | 1 |
| lorazepam | 0.333333 |
| loxapine | 2 |
| lumiracoxib | 1 |
| maprotiline | 1.5 |
| meclizine | 2.25 |
| mesoridazine | 2 |
| metformin | 0.375 |
| methadone | 1.5 |
| methocarbamol | 1.25 |
| methotrexate | 0.5 |
| methscopolamine | 3 |
| methylprednisolone | 0.333333 |
| metoclopramide | 0.4 |
| metoprolol | 0.333333 |
| midazolam | 0.5 |
| mirtazapine | 0.75 |
| molindone | 1 |
| morphine | 0.5 |
| nalbuphine | 1 |
| naratriptan | 1 |
| nefazodone | 0.5 |
| nefopam | 2 |
| neomycin | 1 |
| nifedipine | 0.428571 |
| nitrazepam | 0.333333 |
| nizatidine | 0.333333 |
| nortriptyline | 2.5 |
| olanzapine | 2.333333 |
| opipramol | 3 |
| orphenadrine | 2.833333 |
| oxazepam | 0.2 |
| oxcarbazepine | 1 |
| oxitropium | 2 |
| oxybutynin | 2.444444 |
| oxycodone | 1 |
| paliperidone | 0.5 |
| pancuronium | 1 |
| paracetamol/codeine | 2 |
| paracetamol/codeine/caffeine | 2 |
| paroxetine | 2 |
| pericyazine | 2.5 |
| perphenazine | 1.857143 |
| pethidine | 1 |
| phenelzine | 0.5 |
| phenindamine | 3 |
| pheniramine | 3 |
| phenobarbital | 1 |
| phenyltoloxamine | 3 |
| phenytoin | 0.25 |
| pimozide | 2 |
| piperacillin | 0.5 |
| pramipexole | 0.333333 |
| prednisolone | 0.333333 |
| prednisone | 0.666667 |
| procainamide | 0.5 |
| prochlorperazine | 1.4 |
| procyclidine | 3 |
| promazine | 2.5 |
| promethazine | 3 |
| propantheline | 3 |
| propiverine | 2.5 |
| propoxyphene | 0.5 |
| protriptyline | 3 |
| pseudoephedrine | 1 |
| pyrilamine | 3 |
| quetiapine | 1.666667 |
| quinidine | 0.666667 |
| ranitidine | 1.285714 |
| reboxetine | 0.5 |
| risperidone | 0.666667 |
| rotigotine | 1 |
| selegiline | 0.25 |
| sertraline | 0.4 |
| solifenacin | 2 |
| sumatriptan | 0.5 |
| temazepam | 0.75 |
| theophylline | 1.428571 |
| thioridazine | 3 |
| thiothixene | 1.5 |
| tiagabine | 1 |
| tiotropium | 0.333333 |
| tizanidine | 1.5 |
| tobramycin | 1 |
| tolterodine | 2.8 |
| topiramate | 0.25 |
| tramadol | 1.25 |
| tranylcypromine | 1 |
| trazodone | 0.6 |
| triamcinolone | 0.5 |
| triamterene | 0.666667 |
| triazolam | 1 |
| trifluoperazine | 2.666667 |
| triflupromazine | 2 |
| trihexyphenidyl | 3 |
| trimethobenzamide | 3 |
| trimipramine | 2.666667 |
| triprolidine | 2.5 |
| tropatepine | 3 |
| trospium | 2.666667 |
| valproate | 0.25 |
| vancomycin | 0.5 |
| venlafaxine | 0.4 |
| warfarin | 0.428571 |
| ziprasidone | 0.333333 |
| zolmitriptan | 0.5 |
| zuclopenthixol | 1 |

**Table S3**: APC-analyses for the basic-adjusted models (above) and the fully-adjusted models (below). Each model assumes that one of the APC-terms is zero. Unstandardized regression coefficients (beta) are reported.

| **Model** | **Age (years)** | | | **Period (months)** | | | **Birth cohort** | | |
| --- | --- | --- | --- | --- | --- | --- | --- | --- | --- |
|  | beta | SD | p | beta | SE | p | beta | SE | p |
| Period-cohort |  | | | 0.0089 | 9.7x10^-6^ | <2.0x10^-16^ | -0.057 | 9.3x10^-5^ | <2.0x10^-16^ |
| Age-cohort | 0.11 | 1.2x10^-4^ | <2.0x10^-16^ |  | | | 0.050 | 1.4x10^-4^ | <2.0x10^-16^ |
| Age-period | 0.057 | 9.3x10^-5^ | <2.0x10^-16^ | 0.0042 | 1.2x10^-5^ | <2.0x10^-16^ |  | | |

| **Model** | **Age (years)** | | | **Period (months)** | | | **Birth cohort** | | |
| --- | --- | --- | --- | --- | --- | --- | --- | --- | --- |
|  | beta | SD | p | beta | SE | p | beta | SE | p |
| Period-cohort |  | | | 0.0093 | 9.5x10^-6^ | <2.0x10^-16^ | -0.065 | 8.9x10^-5^ | <2.0x10^-16^ |
| Age-cohort | 0.11 | 1.1x10^-4^ | <2.0x10^-16^ |  | | | 0.046 | 1.4x10^-4^ | <2.0x10^-16^ |
| Age-period | 0.065 | 8.9x10^-5^ | <2.0x10^-16^ | 0.0039 | 1.1x10^-5^ | <2.0x10^-16^ |  | | |

**Table S4**: APC analysis with the monthly number of prescribed anticholinergic drugs as the outcome. Each model assumes that one of the APC-terms is zero. Unstandardized regression coefficients (beta) are reported.

| **Model** | **Age (years)** | | | **Period (months)** | | | **Birth cohort** | | |
| --- | --- | --- | --- | --- | --- | --- | --- | --- | --- |
|  | beta | SD | p | beta | SE | p | beta | SE | p |
| Period-cohort |  | | | 0.0095 | 9.2x10^-6^ | <2.0x10^-16^ | -0.067 | 8.5x10^-5^ | <2.0x10^-16^ |
| Age-cohort | 0.11 | 1.1x10^-4^ | <2.0x10^-16^ |  | | | 0.047 | 1.3x10^-4^ | <2.0x10^-16^ |
| Age-period | 0.067 | 8.6x10^-5^ | <2.0x10^-16^ | 0.0039 | 1.1x10^-5^ | <2.0x10^-16^ |  | | |

**Table S5**: APC-analyses for the basic models, with total number of prescribed drugs as a covariate. Each model assumes that one of the APC-terms is zero. Unstandardized regression coefficients (beta) are reported.

| **Model** | **Age (years)** | | | **Period (months)** | | | **Birth cohort** | | |
| --- | --- | --- | --- | --- | --- | --- | --- | --- | --- |
|  | beta | SD | p | beta | SE | p | beta | SE | p |
| Period-cohort |  | | | 2.2x10^-4^ | 6.9x10^-6^ | <2.0x10^-16^ | 0.0029 | 6.4x10^-6^ | <2.0x10^-16^ |
| Age-cohort | 0.0027 | 8.3x10^-5^ | <2.0x10^-16^ |  | | | 0.0055 | 1.0x10^-4^ | <2.0x10^-16^ |
| Age-period | -0.0029 | 6.4x10^-5^ | <2.0x10^-16^ | 4.6x10^-4^ | 8.3x10^-6^ | <2.0x10^-16^ |  | | |

**Table S6**: Results of the models predicting monthly anticholinergic burden due to different drug classes as a function of deprivation, smoking, BMI, sex, education, region, alcohol consumption, physical activity, and age.

| **Antidepressant** | | | | |
| --- | --- | --- | --- | --- |
| **Predictor** | **Level** | **Beta** | **SE** | **p** |
| Deprivation |  | 0.002 | 3.2x10^-4^ | <2.0x10^-16^ |
| Smoking (*ref: non-smoker*) | Previous smoker | 0.039 | 0.0020 | 9.5x10^-9^ |
|  | Current smoker | 0.099 | 0.0032 | <2.0x10^-16^ |
| BMI |  | 0.004 | 2.0x10^-4^ | <2.0x10^-16^ |
| Sex | Male | -0.117 | 0.0019 | <2.0x10^-16^ |
| Education | graduate degree | -0.045 | 0.0020 | <2.0x10^-16^ |
| Region (*ref: England*) | Scotland | -0.033 | 0.0032 | <2.0x10^-16^ |
|  | Wales | 0.020 | 0.0032 | <2.0x10^-16^ |
| Alcohol consumption (*ref: daily or almost daily consumption*) | Three or four times a week | -0.011 | 0.0028 | 7.5x10^-10^ |
|  | Once or twice a week | 0.007 | 0.0028 | 6.3x10^-5^ |
|  | Once to thrice a month | 0.026 | 0.0034 | 1.1x10^-2^ |
|  | Special occasions only | 0.044 | 0.0035 | 3.2x10^-14^ |
|  | Never | 0.060 | 0.0040 | <2.0x10^-16^ |
| Physical activity (*ref: mild or no physical activity*) | Moderate | -0.031 | 0.0021 | <2.0x10^-16^ |
|  | Strenuous | -0.073 | 0.0036 | <2.0x10^-16^ |

| **Acid disorders** | | | | |
| --- | --- | --- | --- | --- |
| **Predictor** | **Level** | **Beta** | **SE** | **p** |
| Deprivation |  | 0.001 | 1.11x10^-4^ | 3.4x10^-6^ |
| Smoking (*ref: non-smoker*) | Previous smoker | 0.011 | 6.87x10^-4^ | <2.0x10^-16^ |
|  | Current smoker | 0.012 | 0.0011 | <2.0x10^-16^ |
| BMI |  | 0.002 | 6.95x10^-5^ | <2.0x10^-16^ |
| Sex | Male | -0.002 | 6.49x10^-4^ | 0.0025 |
| Education | graduate degree | -0.020 | 7.00x10^-4^ | <2.0x10^-16^ |
| Region (*ref: England*) | Scotland | -0.024 | 0.0011 | <2.0x10^-16^ |
|  | Wales | 0.005 | 0.0011 | 1.9x10^-6^ |
| Alcohol consumption (*ref: daily or almost daily consumption*) | Three or four times a week | -2.0x10^-4^ | 9.5x10^-4^ | 0.83 |
|  | Once or twice a week | 0.005 | 9.36x10^-4^ | 6.1x10^-9^ |
|  | Once to thrice a month | 0.008 | 0.0012 | 1.3 x10^-12^ |
|  | Special occasions only | 0.013 | 0.0012 | <2.0x10^-16^ |
|  | Never | 0.022 | 0.0014 | <2.0x10^-16^ |
| Physical activity (*ref: mild or no physical activity*) | Moderate | -0.008 | 7.27x10^-4^ | <2.0x10^-16^ |
|  | Strenuous | -0.018 | 0.0012 | <2.0x10^-16^ |

| **Analgesic** | | | | |
| --- | --- | --- | --- | --- |
| **Predictor** | **Level** | **Beta** | **SE** | **p** |
| Deprivation |  | 0.004 | 1.77x10^-4^ | <2.0x10^-16^ |
| Smoking (*ref: non-smoker*) | Previous smoker | 0.023 | 0.0011 | <2.0x10^-16^ |
|  | Current smoker | 0.048 | 0.0018 | <2.0x10^-16^ |
| BMI |  | 0.006 | 1.12x10^-4^ | <2.0x10^-16^ |
| Sex | Male | -0.028 | 0.0011 | <2.0x10^-16^ |
| Education | graduate degree | -0.044 | 0.0011 | <2.0x10^-16^ |
| Region (*ref: England*) | Scotland | 0.036 | 0.0016 | <2.0x10^-16^ |
|  | Wales | -0.014 | 0.0018 | 2.9x10^-15^ |
| Alcohol consumption (*ref: daily or almost daily consumption*) | Three or four times a week | -2.47x10^-4^ | 0.0016 | 0.87 |
|  | Once or twice a week | 0.011 | 0.0015 | 1.5x10^-13^ |
|  | Once to thrice a month | 0.020 | 0.0019 | <2.0x10^-16^ |
|  | Special occasions only | 0.033 | 0.0019 | <2.0x10^-16^ |
|  | Never | 0.043 | 0.0022 | <2.0x10^-16^ |
| Physical activity (*ref: mild or no physical activity*) | Moderate | -0.015 | 0.0012 | <2.0x10^-16^ |
|  | Strenuous | -0.035 | 0.0020 | <2.0x10^-16^ |

| **Antithrombotic** | | | | |
| --- | --- | --- | --- | --- |
| **Predictor** | **Level** | **Beta** | **SE** | **p** |
| Deprivation |  | 4.1x10^-4^ | 5.3x10^-4^ | 0.44 |
| Smoking (*ref: non-smoker*) | Previous smoker | 0.014 | 0.0032 | 1.7x10^-5^ |
|  | Current smoker | 0.005 | 0.0055 | 0.327 |
| BMI |  | 0.008 | 3.3x10^-4^ | <2.0x10^-16^ |
| Sex | Male | 0.104 | 0.0033 | <2.0x10^-16^ |
| Education | graduate degree | -0.007 | 0.0033 | 0.034 |
| Region (*ref: England*) | Scotland | -0.011 | 0.0052 | 0.039 |
|  | Wales | 0.033 | 0.0049 | 1.0x10^-11^ |
| Alcohol consumption (*ref: daily or almost daily consumption*) | Three or four times a week | -0.011 | 0.0044 | 0.016 |
|  | Once or twice a week | -0.006 | 0.0043 | 0.139 |
|  | Once to thrice a month | -1.4x10^-4^ | 0.0057 | 0.980 |
|  | Special occasions only | 0.011 | 0.0057 | 0.047 |
|  | Never | 0.026 | 0.0063 | 3.5x10^-5^ |
| Physical activity (*ref: mild or no physical activity*) | Moderate | -0.017 | 0.0034 | 9.0x10^-7^ |
|  | Strenuous | -0.026 | 0.0061 | 2.9x10^-5^ |

| **Diabetes** | | | | |
| --- | --- | --- | --- | --- |
| **Predictor** | **Level** | **Beta** | **SE** | **p** |
| Deprivation |  | 0.006 | 4.2x10^-4^ | <2.0x10^-16^ |
| Smoking (*ref: non-smoker*) | Previous smoker | 0.020 | 0.0027 | 2.7x10^-13^ |
|  | Current smoker | 0.024 | 0.0043 | 4.2x10^-8^ |
| BMI |  | 0.019 | 2.9x10^-4^ | <2.0x10^-16^ |
| Sex | Male | 0.111 | 0.0028 | <2.0x10^-16^ |
| Education | graduate degree | -0.009 | 0.0028 | 0.001 |
| Region (*ref: England*) | Scotland | -0.015 | 0.0043 | 0.001 |
|  | Wales | 0.011 | 0.0042 | 0.009 |
| Alcohol consumption (*ref: daily or almost daily consumption*) | Three or four times a week | 0.010 | 0.0041 | 0.013 |
|  | Once or twice a week | 0.042 | 0.0039 | <2.0x10^-16^ |
|  | Once to thrice a month | 0.068 | 0.0047 | <2.0x10^-16^ |
|  | Special occasions only | 0.100 | 0.0047 | <2.0x10^-16^ |
|  | Never | 0.129 | 0.0050 | <2.0x10^-16^ |
| Physical activity (*ref: mild or no physical activity*) | Moderate | -0.034 | 0.0027 | <2.0x10^-16^ |
|  | Strenuous | -0.088 | 0.0057 | <2.0x10^-16^ |

| **Cardiovascular** | | | | |
| --- | --- | --- | --- | --- |
| **Predictor** | **Level** | **Beta** | **SE** | **p** |
| Deprivation |  | 0.002 | 3.1x10^-4^ | 4.1x10^-16^ |
| Smoking (*ref: non-smoker*) | Previous smoker | 0.020 | 0.0019 | <2.0x10^-16^ |
|  | Current smoker | 0.017 | 0.0031 | 1.0x10^-7^ |
| BMI |  | 0.012 | 1.9x10^-4^ | <2.0x10^-16^ |
| Sex | Male | 0.030 | 0.0018 | <2.0x10^-16^ |
| Education | graduate degree | -0.035 | 0.0020 | <2.0x10^-16^ |
| Region (*ref: England*) | Scotland | 3.3x10^-4^ | 0.0029 | 0.91 |
|  | Wales | 0.013 | 0.0030 | 2.1x10^-5^ |
| Alcohol consumption (*ref: daily or almost daily consumption*) | Three or four times a week | -0.001 | 0.0026 | 0.61 |
|  | Once or twice a week | 0.004 | 0.0026 | 0.14 |
|  | Once to thrice a month | 0.008 | 0.0033 | 0.02 |
|  | Special occasions only | 0.029 | 0.0033 | <2.0x10^-16^ |
|  | Never | 0.038 | 0.0037 | <2.0x10^-16^ |
| Physical activity (*ref: mild or no physical activity*) | Moderate | -0.029 | 0.0020 | <2.0x10^-16^ |
|  | Strenuous | -0.070 | 0.0038 | <2.0x10^-16^ |

| **Gastrointestinal** | | | | |
| --- | --- | --- | --- | --- |
| **Predictor** | **Level** | **Beta** | **SE** | **p** |
| Deprivation |  | 0.001 | 1.1x10^-4^ | 3.9x10^-14^ |
| Smoking (*ref: non-smoker*) | Previous smoker | 0.006 | 7.2x10^-4^ | 2.9x10^-15^ |
|  | Current smoker | 0.007 | 0.0012 | 1.1x10^-9^ |
| BMI |  | 3.3x10^-4^ | 7.2x10^-5^ | 5.9x10^-6^ |
| Sex | Male | -0.025 | 7.0x10^-4^ | <2.0x10^-16^ |
| Education | graduate degree | -0.013 | 7.4x10^-4^ | <2.0x10^-16^ |
| Region (*ref: England*) | Scotland | -0.007 | 0.0011 | 2.3x10^-10^ |
|  | Wales | 0.002 | 0.0011 | 0.03 |
| Alcohol consumption (*ref: daily or almost daily consumption*) | Three or four times a week | -0.001 | 0.0010 | 0.46 |
|  | Once or twice a week | 0.006 | 0.0010 | 7.0x10^-9^ |
|  | Once to thrice a month | 0.009 | 0.0012 | 1.2x10^-12^ |
|  | Special occasions only | 0.015 | 0.0012 | <2.0x10^-16^ |
|  | Never | 0.021 | 0.0014 | <2.0x10^-16^ |
| Physical activity (*ref: mild or no physical activity*) | Moderate | -0.008 | 7.5x10^-4^ | <2.0x10^-16^ |
|  | Strenuous | -0.015 | 0.0013 | <2.0x10^-16^ |

| **Psycholeptic** | | | | |
| --- | --- | --- | --- | --- |
| **Predictor** | **Level** | **Beta** | **SE** | **p** |
| Deprivation |  | 0.001 | 1.0x10^-4^ | <2.0x10^-16^ |
| Smoking (*ref: non-smoker*) | Previous smoker | 0.007 | 6.3x10^-4^ | <2.0x10^-16^ |
|  | Current smoker | 0.015 | 0.0010 | <2.0x10^-16^ |
| BMI |  | 0.001 | 6.4x10^-5^ | <2.0x10^-16^ |
| Sex | Male | -0.027 | 6.1x10^-4^ | <2.0x10^-16^ |
| Education | graduate degree | -0.007 | 6.3x10^-4^ | <2.0x10^-16^ |
| Region (*ref: England*) | Scotland | -0.005 | 0.0010 | 1.6x10^-6^ |
|  | Wales | 0.006 | 0.0010 | 3.8x10^-10^ |
| Alcohol consumption (*ref: daily or almost daily consumption*) | Three or four times a week | -0.004 | 8.8x10^-4^ | 6.2x10^-6^ |
|  | Once or twice a week | -0.001 | 8.6x10^-4^ | 0.48 |
|  | Once to thrice a month | 0.004 | 0.0011 | 4.1x10^-5^ |
|  | Special occasions only | 0.009 | 0.0011 | 1.5x10^-15^ |
|  | Never | 0.017 | 0.0012 | <2.0x10^-16^ |
| Physical activity (*ref: mild or no physical activity*) | Moderate | -0.007 | 6.7x10^-4^ | <2.0x10^-16^ |
|  | Strenuous | -0.013 | 0.0011 | <2.0x10^-16^ |

| **Respiratory** | | | | |
| --- | --- | --- | --- | --- |
| **Predictor** | **Level** | **Beta** | **SE** | **p** |
| Deprivation |  | 0.002 | 1.2x10^-4^ | <2.0x10^-16^ |
| Smoking (*ref: non-smoker*) | Previous smoker | 0.010 | 7.7x10^-4^ | <2.0x10^-16^ |
|  | Current smoker | 0.015 | 0.0012 | <2.0x10^-16^ |
| BMI |  | 0.002 | 7.7x10^-5^ | <2.0x10^-16^ |
| Sex | Male | -0.025 | 7.3x10^-4^ | <2.0x10^-16^ |
| Education | graduate degree | -0.015 | 7.7x10^-4^ | <2.0x10^-16^ |
| Region (*ref: England*) | Scotland | -0.018 | 0.0012 | <2.0x10^-16^ |
|  | Wales | 0.030 | 0.0012 | <2.0x10^-16^ |
| Alcohol consumption (*ref: daily or almost daily consumption*) | Three or four times a week | -0.002 | 0.0011 | 0.074 |
|  | Once or twice a week | 0.003 | 0.0010 | 0.006 |
|  | Once to thrice a month | 0.004 | 0.0013 | 6.8x10^-4^ |
|  | Special occasions only | 0.013 | 0.0013 | <2.0x10^-16^ |
|  | Never | 0.023 | 0.0015 | <2.0x10^-16^ |
| Physical activity (*ref: mild or no physical activity*) | Moderate | -0.007 | 8.2x10^-4^ | <2.0x10^-16^ |
|  | Strenuous | -0.015 | 0.0013 | <2.0x10^-16^ |

| **Urological** | | | | |
| --- | --- | --- | --- | --- |
| **Predictor** | **Level** | **Beta** | **SE** | **p** |
| Deprivation |  | 0.003 | 7.2x10^-4^ | 2.9x10^-5^ |
| Smoking (*ref: non-smoker*) | Previous smoker | 0.023 | 0.0044 | 1.3x10^-7^ |
|  | Current smoker | 0.008 | 0.0075 | 0.31 |
| BMI |  | 0.007 | 4.4x10^-4^ | <2.0x10^-16^ |
| Sex | Male | -0.081 | 0.0043 | <2.0x10^-16^ |
| Education | graduate degree | -0.038 | 0.0046 | <2.0x10^-16^ |
| Region (*ref: England*) | Scotland | 0.019 | 0.0067 | 0.005 |
|  | Wales | 0.003 | 0.0072 | 0.70 |
| Alcohol consumption (*ref: daily or almost daily consumption*) | Three or four times a week | 0.015 | 0.0064 | 0.02 |
|  | Once or twice a week | 0.045 | 0.0062 | 6.1x10^-13^ |
|  | Once to thrice a month | 0.060 | 0.0077 | 3.3x10^-15^ |
|  | Special occasions only | 0.080 | 0.0077 | <2.0x10^-16^ |
|  | Never | 0.090 | 0.0086 | <2.0x10^-16^ |
| Physical activity (*ref: mild or no physical activity*) | Moderate | -0.007 | 0.0047 | 0.12 |
|  | Strenuous | -0.029 | 0.0084 | 4.0x10^-4^ |

| **Other** | | | | |
| --- | --- | --- | --- | --- |
| **Predictor** | **Level** | **Beta** | **SE** | **p** |
| Deprivation |  | 2.0x10^-4^ | 6.4x10^-5^ | 0.001 |
| Smoking (*ref: non-smoker*) | Previous smoker | 0.004 | 3.9x10^-4^ | <2.0x10^-16^ |
|  | Current smoker | 0.006 | 6.4x10^-4^ | <2.0x10^-16^ |
| BMI |  | 0.001 | 4.0x10^-5^ | <2.0x10^-16^ |
| Sex | Male | -0.002 | 3.7x10^-4^ | 1.2x10^-6^ |
| Education | graduate degree | -0.006 | 3.9x10^-4^ | <2.0x10^-16^ |
| Region (*ref: England*) | Scotland | -0.006 | 6.1x10^-4^ | <2.0x10^-16^ |
|  | Wales | 0.004 | 6.2x10^-4^ | 2.9x10^-9^ |
| Alcohol consumption (*ref: daily or almost daily consumption*) | Three or four times a week | -0.001 | 5.4x10^-4^ | 0.15 |
|  | Once or twice a week | 0.001 | 5.3x10^-4^ | 0.039 |
|  | Once to thrice a month | 0.002 | 6.7x10^-4^ | 0.011 |
|  | Special occasions only | 0.005 | 6.9x10^-4^ | 6.4x10^-15^ |
|  | Never | 0.012 | 7.9x10^-4^ | <2.0x10^-16^ |
| Physical activity (*ref: mild or no physical activity*) | Moderate | -0.004 | 4.2x10^-4^ | <2.0x10^-16^ |
|  | Strenuous | -0.007 | 6.8x10^-4^ | <2.0x10^-16^ |

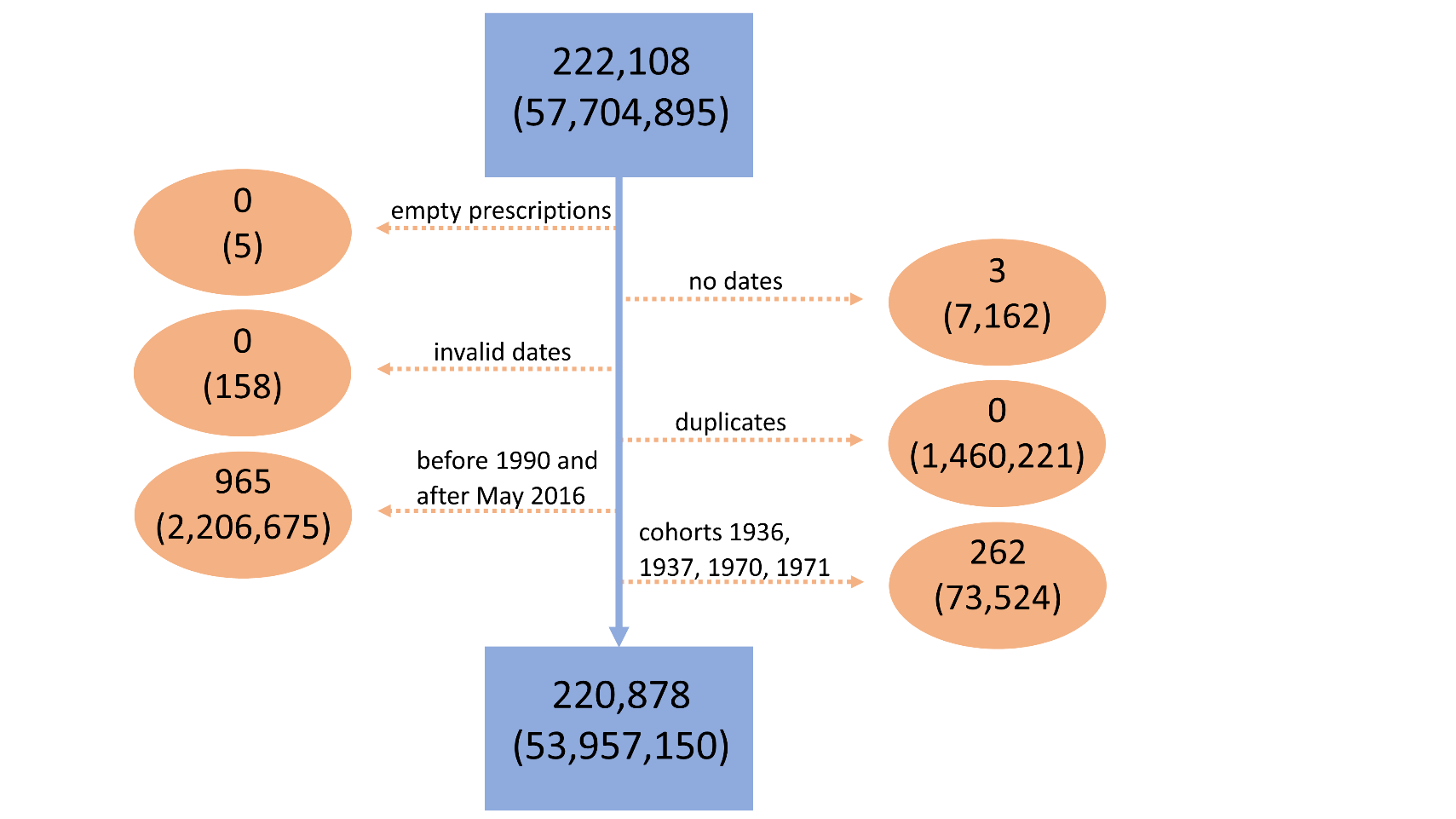

**Figure S1**: Prescriptions without drug and Read-code were removed. We removed prescriptions without recorded dates, prescriptions dated before or at participants’ dates of birth or after the participants’ dates of death, and duplicate prescriptions (= the same prescription issued to the same individual on the same day). We removed prescriptions issued after the date on which sampling was terminated for any data provider and all months for which the number of recorded participants was lower than 10% of the monthly median over the entire sampling period (median=20,647), resulting in the removal of prescriptions prior to and including December 1989 and those after and including June 2016. For mixed-models analyses, we additionally removed birth cohorts 1936, 1937, 1970, and 1971 due to low numbers of individuals (1, 182, 81, 1, respectively; median=6,487). The final sample consisted of 53,956,916 prescriptions issued to 220,867 individuals.

Depiction of the data cleaning procedure. The top numbers refer to the numbers of participants, the lower numbers in brackets refer to the numbers of prescriptions in the sample. The top and bottom blue box indicate the numbers of participants/prescriptions before and after data cleaning, respectively. Each orange oval represents a step in data cleaning; the steps were performed sequentially from top to the bottom.

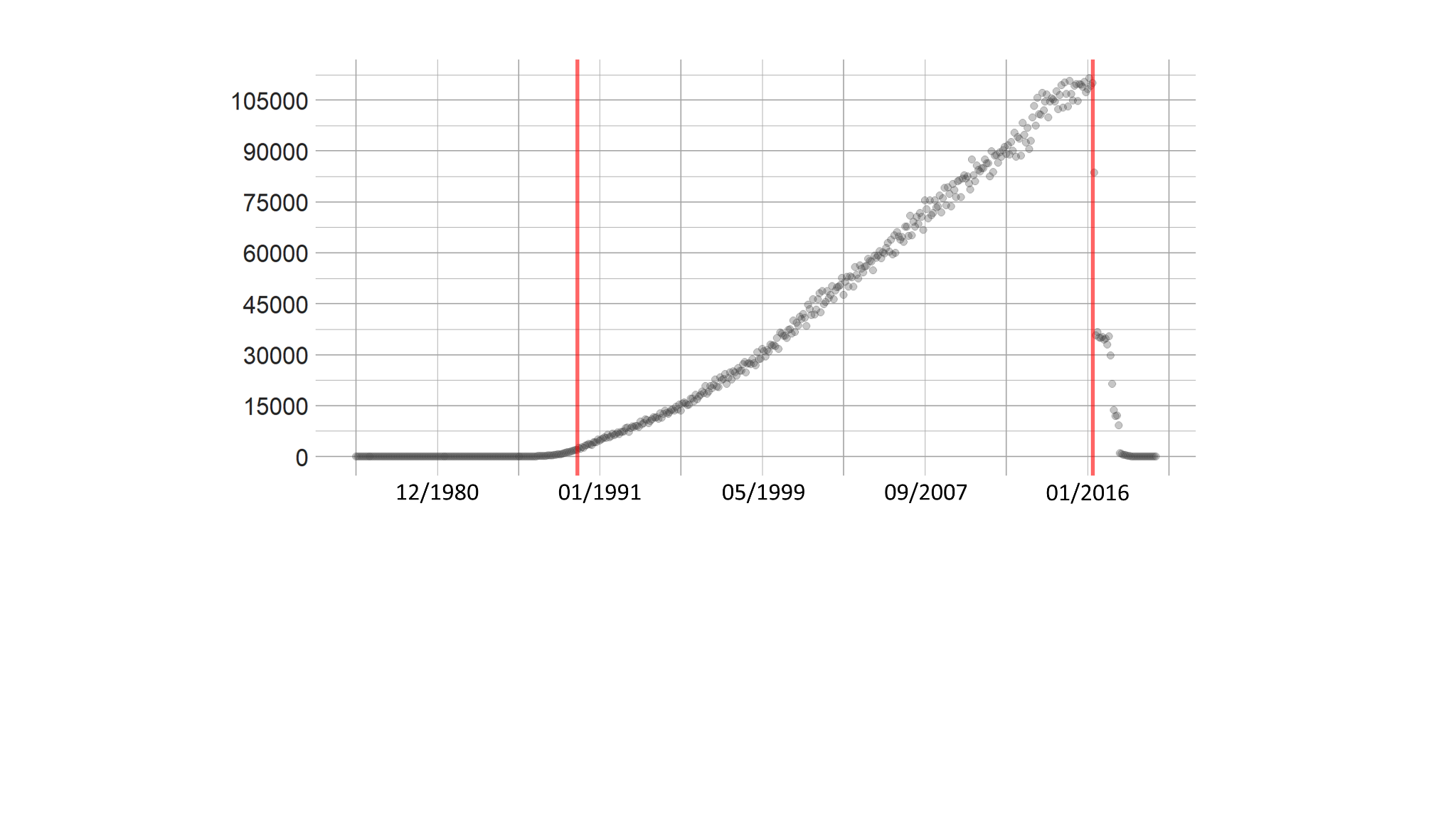

**Figure S2**: The removal of the period prior to and including December 1989 and after and including June 2016. The x-axis represents months in the original dataset, the y axis represents the number of participants that were issued a prescription each month. The vertical red lines indicate December 1989 and June 2016; the period between those dates was retained for further analysis.

**Figure S3**: In the raw data, the unit of observation was a prescription. For longitudinal analyses, the unit of observation was transformed into a participant-period combination, with the anticholinergic burden for each participant the sum over the anticholinergic burden of all prescriptions for that participant in that period. Monthly anticholinergic burden was the outcome for all models except mixed-effects models, where – for computational parsimony – it was yearly anticholinergic burden. For analyses of lifestyle- and demographic factors, the data were transformed so that individual participants were units of observations. Person-time was calculated by subtracting the date of the participant’s appearance in the dataset from (1) the participant’s date of death or (2) the last date in the dataset (whichever came first). For each participant we then summed their anticholinergic burden across their prescriptions and divided it by their person-time. We removed participants with a person-time of <12 months (n=11).
An example illustrating the data transformation process and the calculations involved is depicted below. The upper table (“prescriptions data frame”) represents the format of the raw data after the computation and addition of anticholinergic scores. Each row is a single prescription and the “AC-burden” is the anticholinergic score associated with a given prescription. The lower left table (“id-period data frame”) represents the format used for the analyses of longitudinal trends; in the example below, the period is year, whereas our analyses – depending on the model – used either year or month. In the id-period data frame, each row is an id-period combination, with the anticholinergic burden (“AC-burden”) the sum of the anticholinergic scores of all prescriptions taken by a participant in a given month. The lower right table (“id data frame”) represents the format used for the analysis of the association between demographic- and lifestyle factors and anticholinergic burden. In this table, each row is a participant and the anticholinergic burden is the average anticholinergic burden in a given period (year in the example below) for that participant. The example below is an imaginary example which assumes that all three participants where part of the sample throughout the entire sampling period.

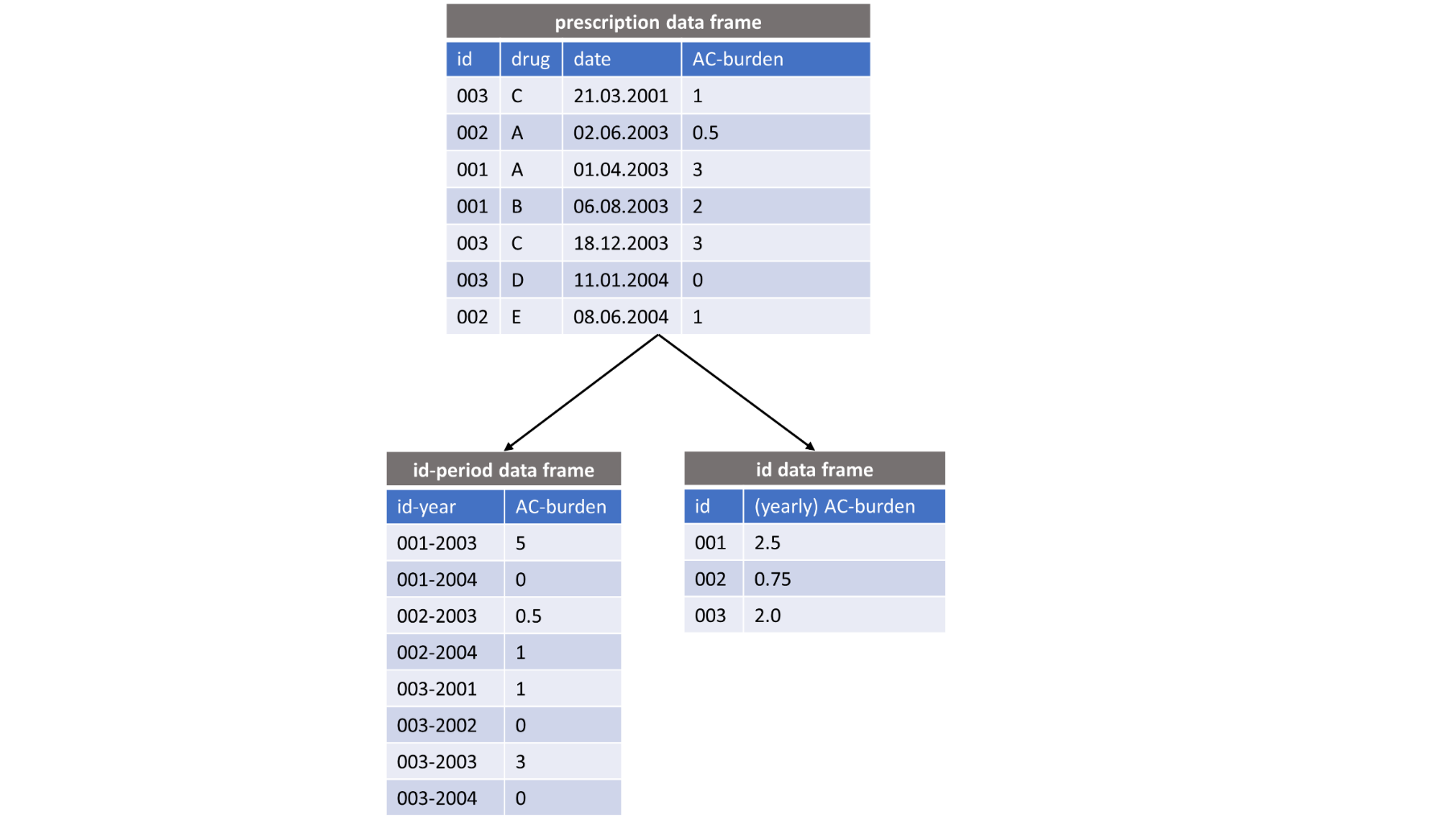

**Figure S4**: Distribution of birth cohorts in the sample.

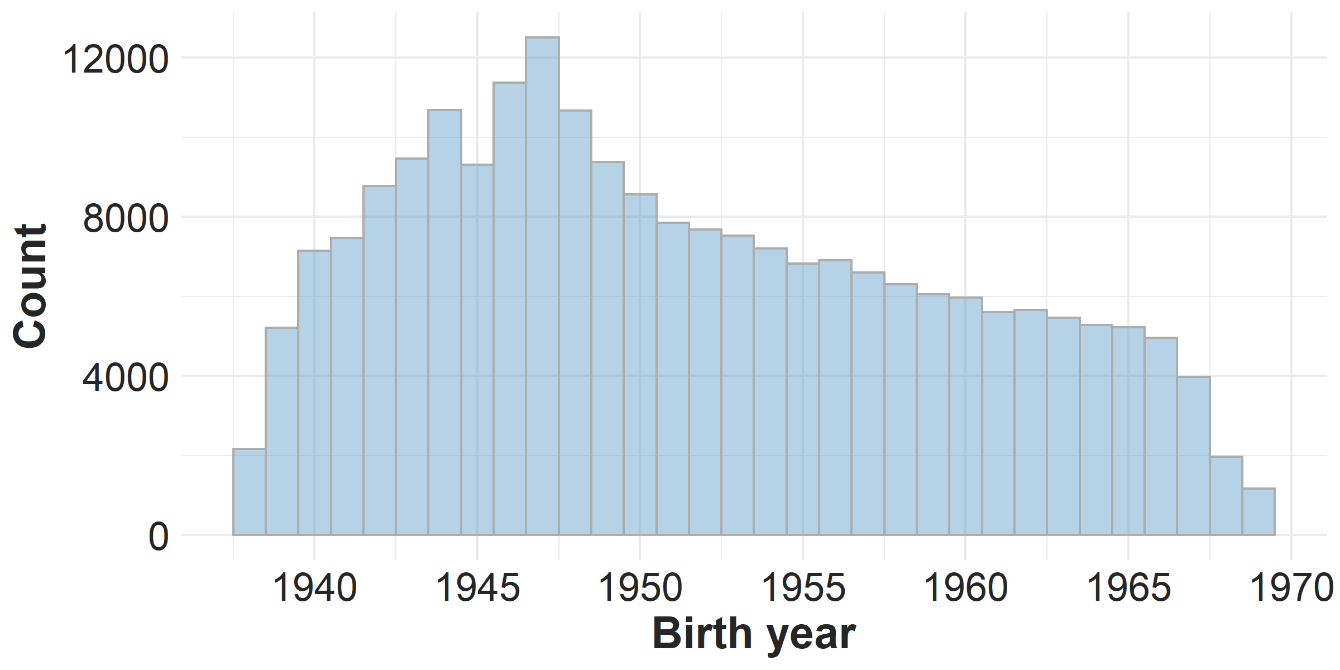

**Figure S5**: Changes in the proportions of anticholinergic burden due to the different categories of drugs (**A**) and in the proportions of the total numbers of prescribed drugs from each category (**B**). The categories refer to mild (greater than 0, equal to or lower than 1), moderate (greater than 1, equal to or lower than 2), and strong (greater than 2) anticholinergic activity.

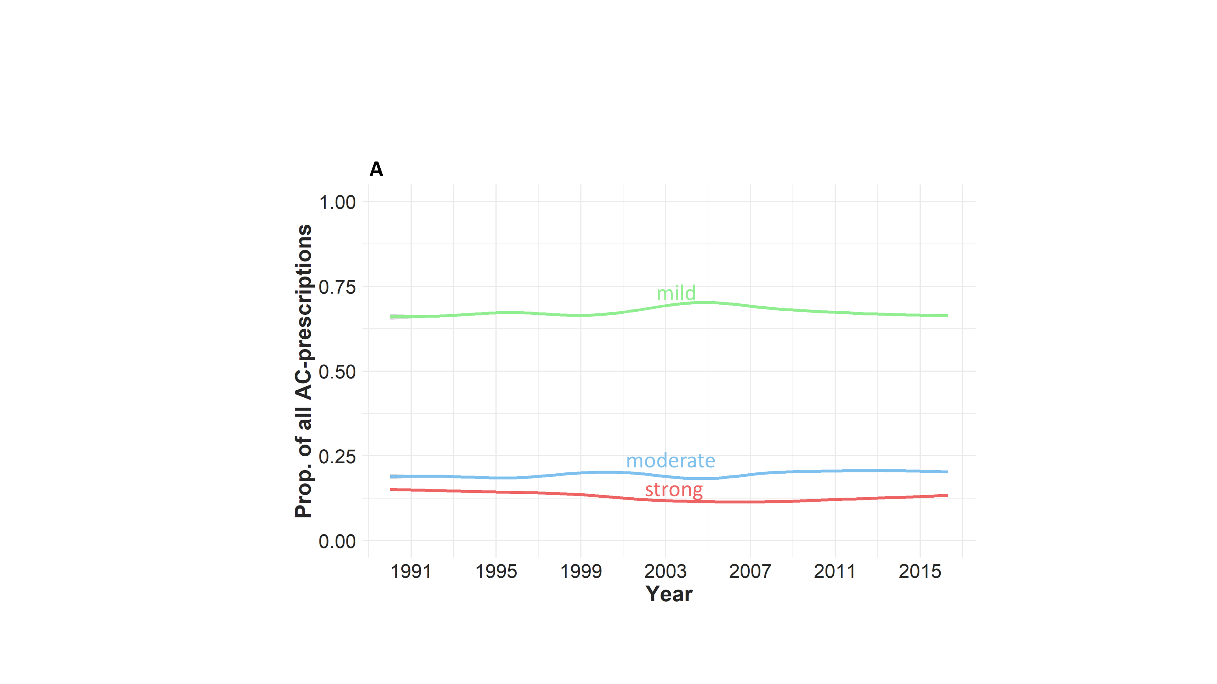

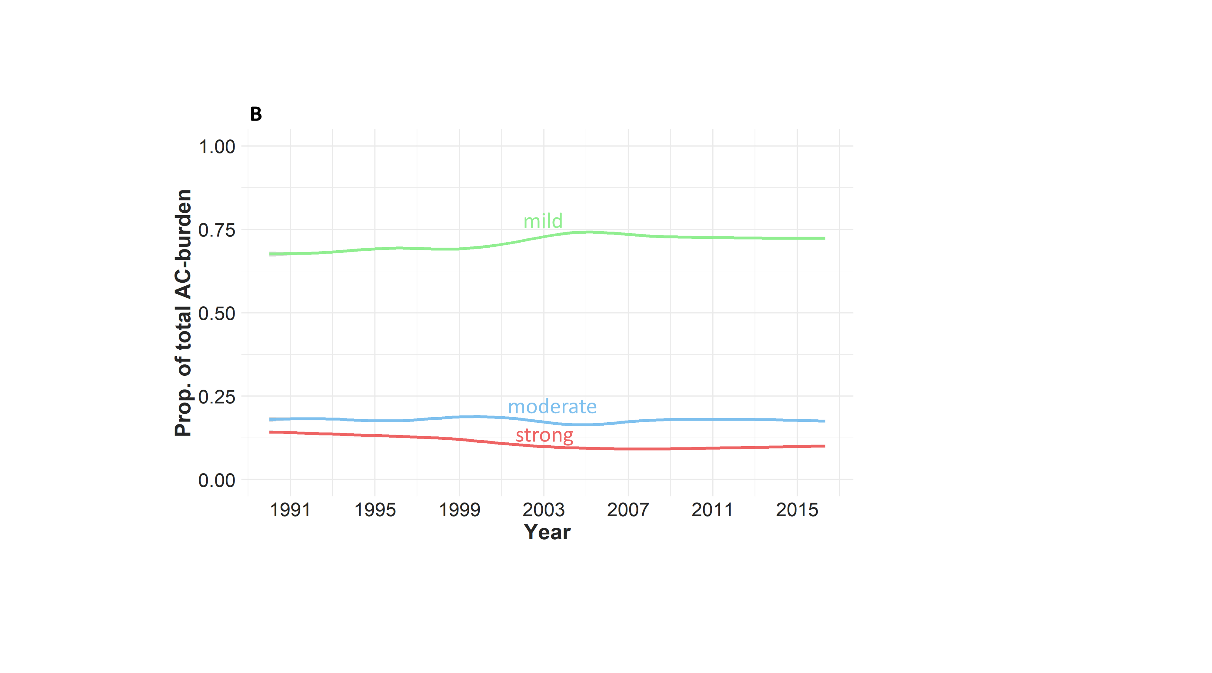

**Figure S6**: Plots of anticholinergic burden for each drug class. Displayed are the period-cohort model with cohort as a random effect (left column), the age-cohort model with cohort as a random effect (middle column), and the age-period model with period as the random effect (right column). To increase accuracy, the plotting was done by using the id-month data frame (with monthly anticholinergic burden as the outcome). The plots were generated using generalised additive model smoothing.

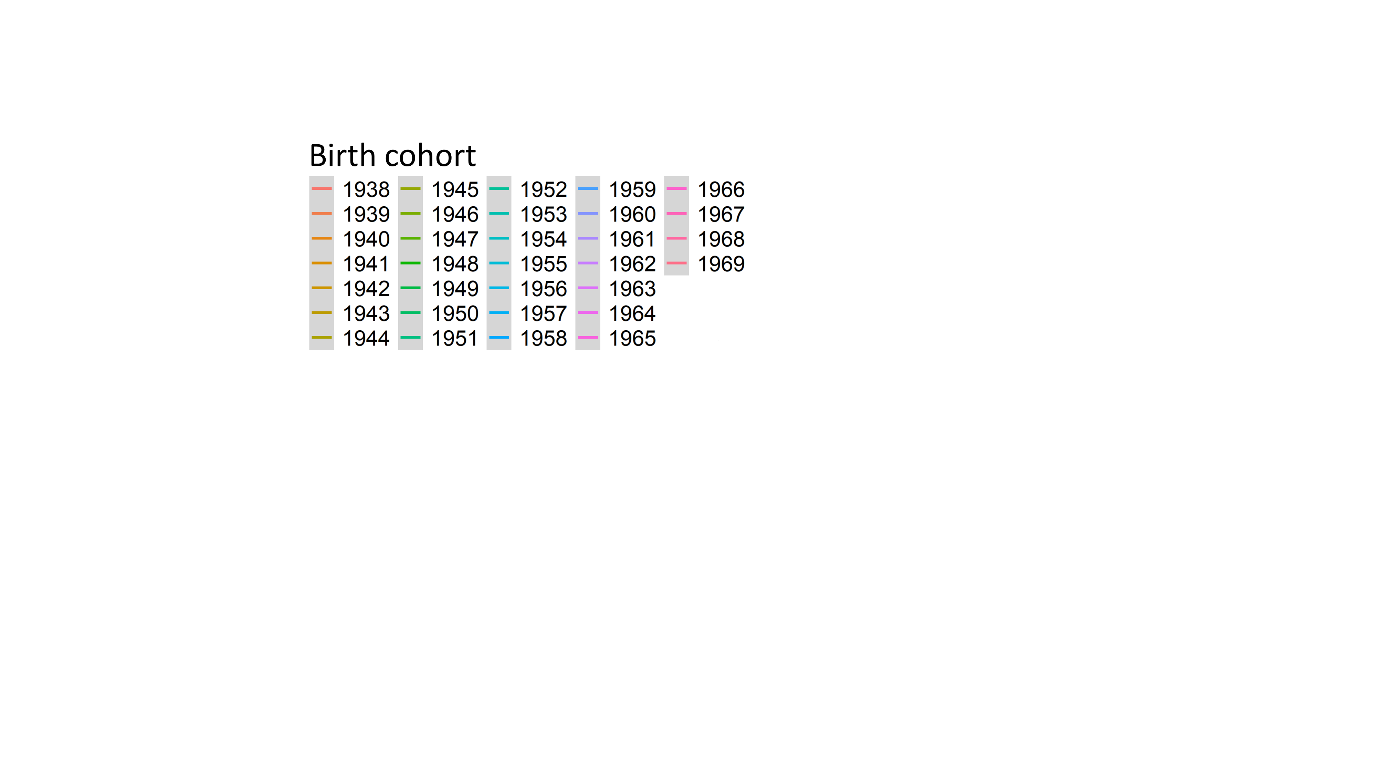

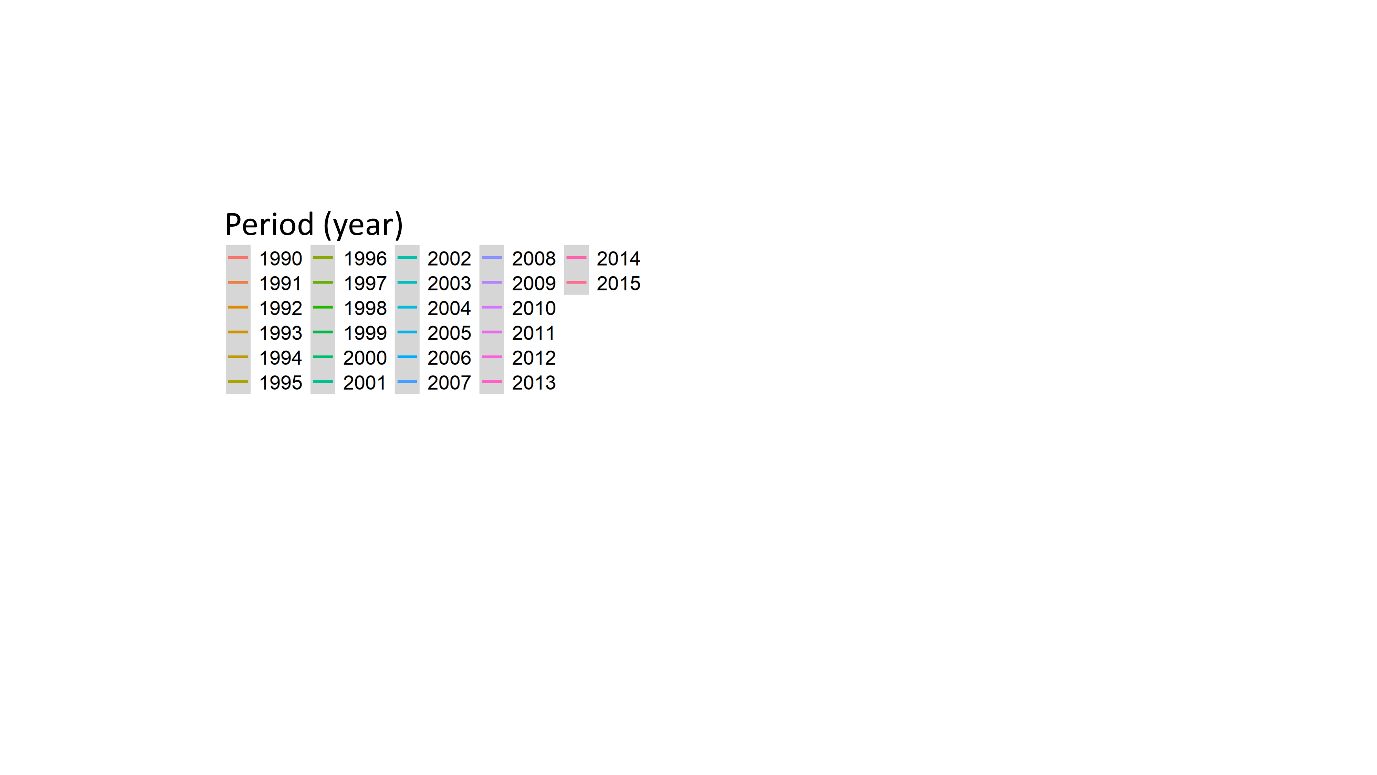

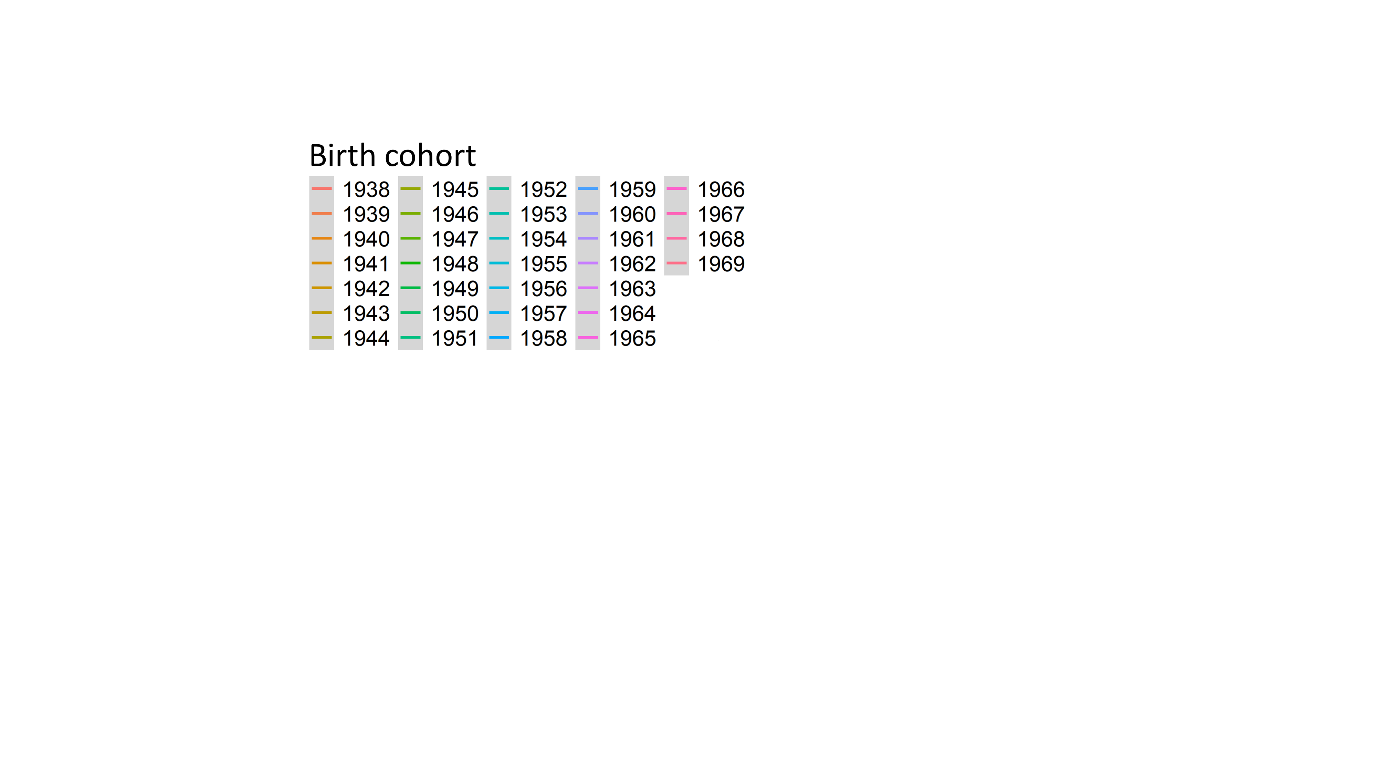

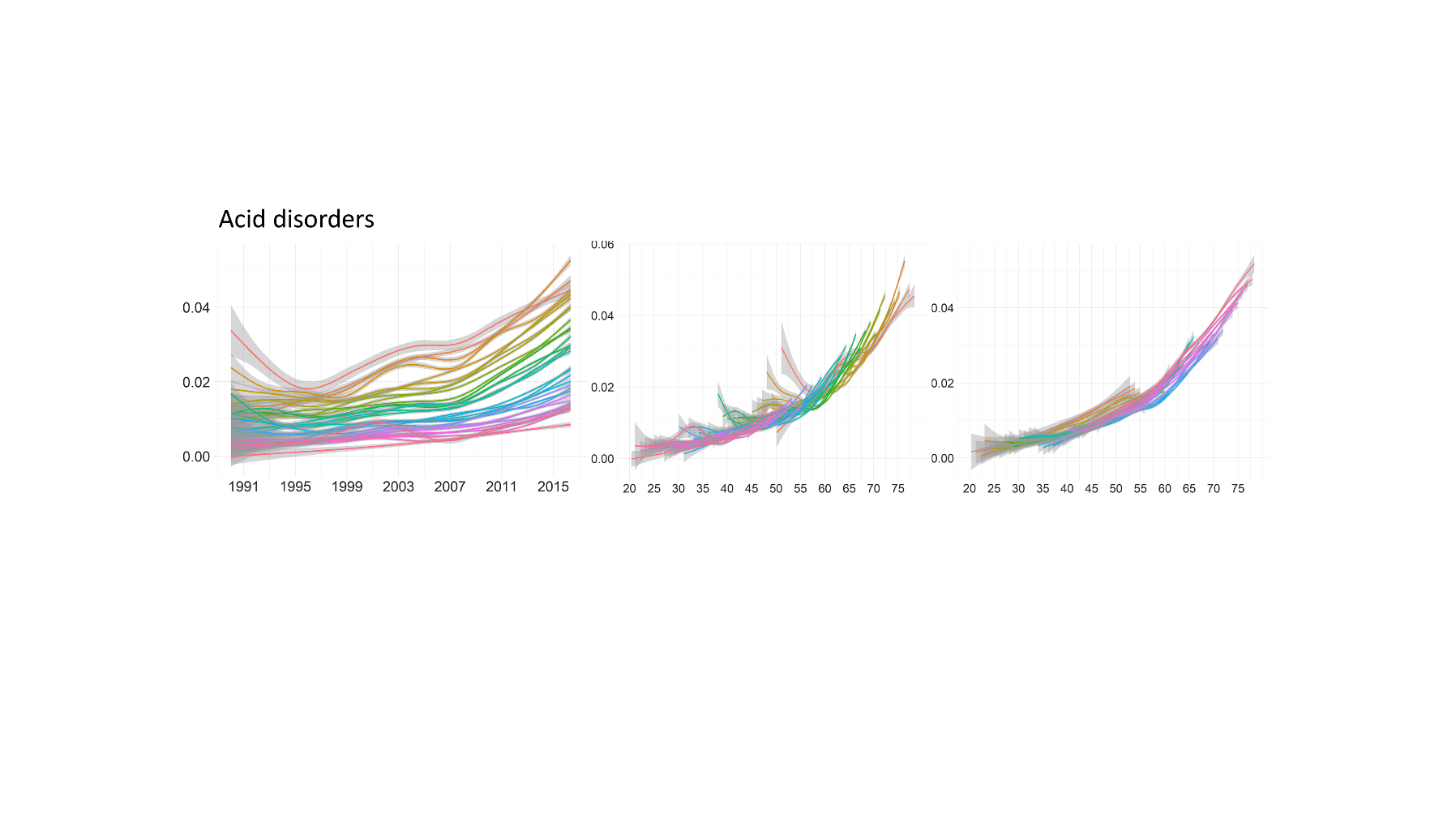

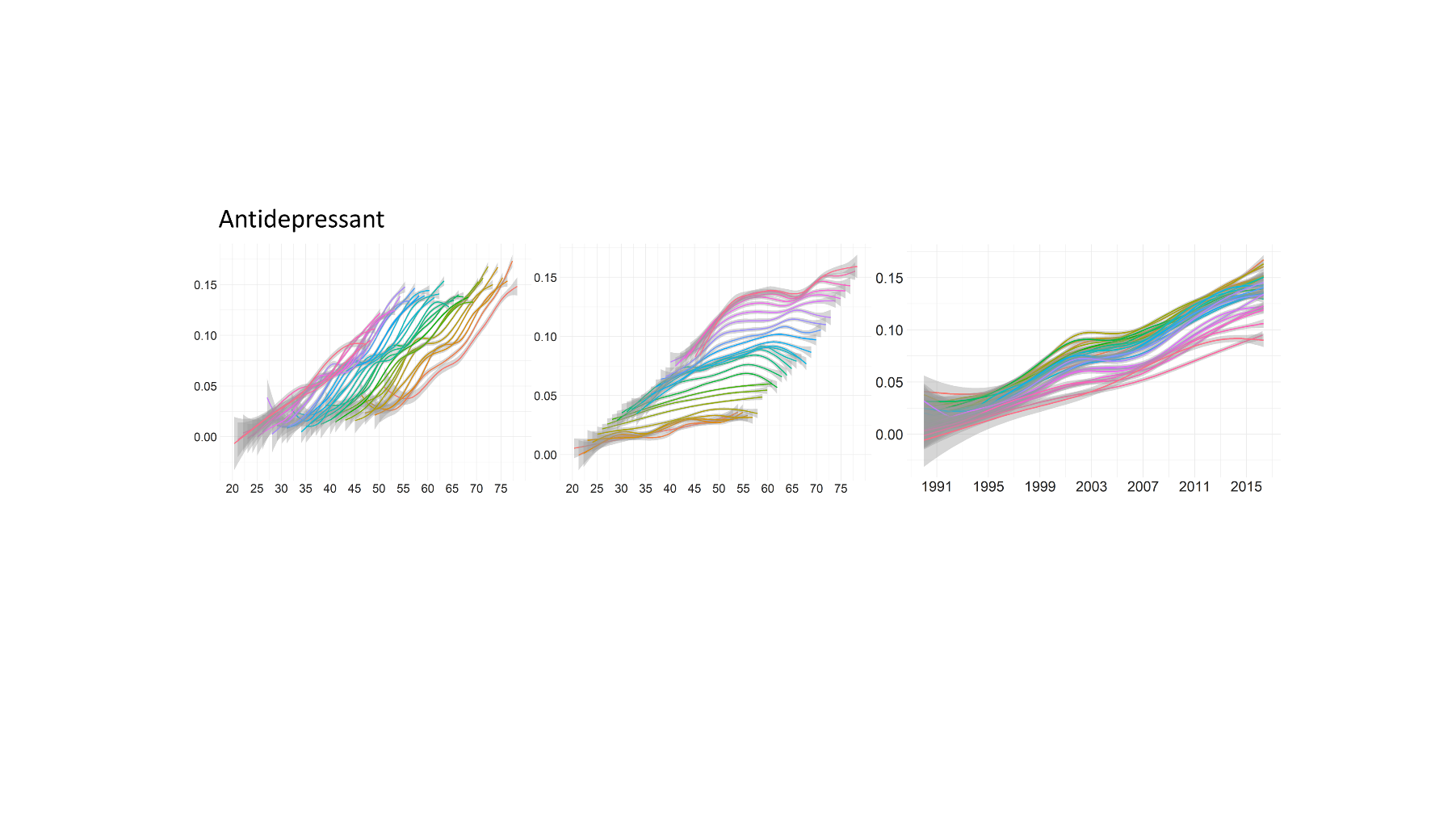

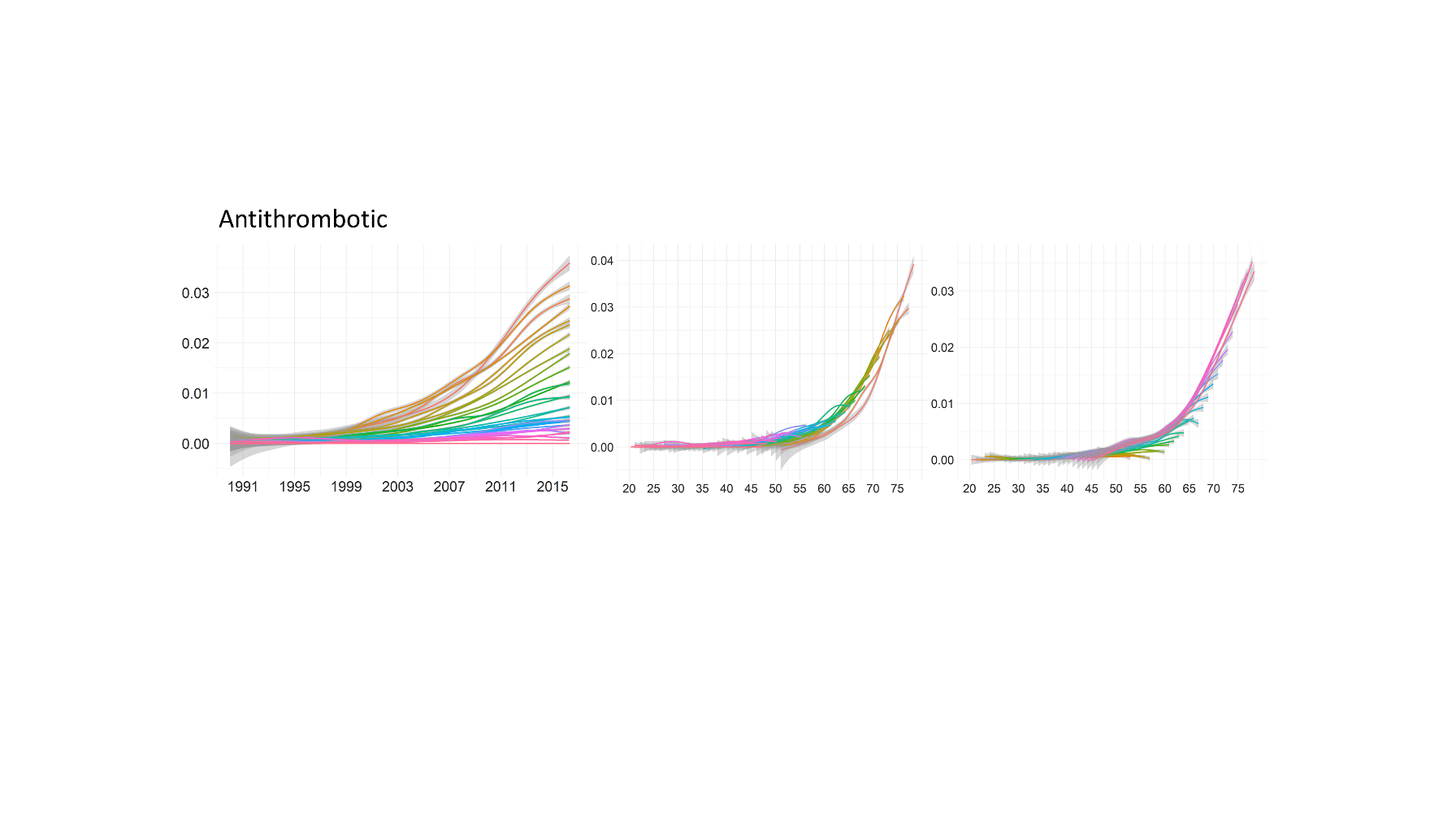

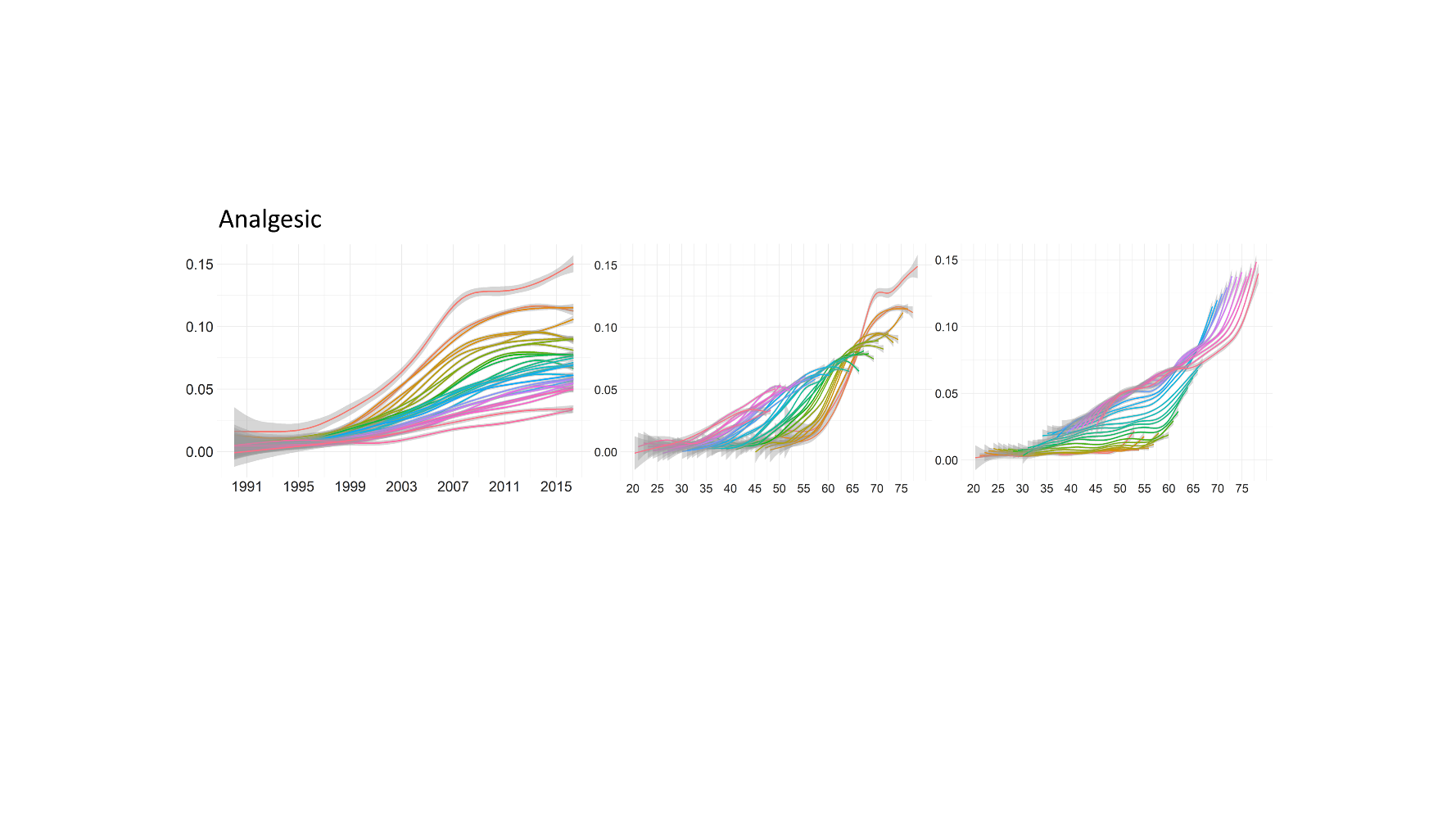

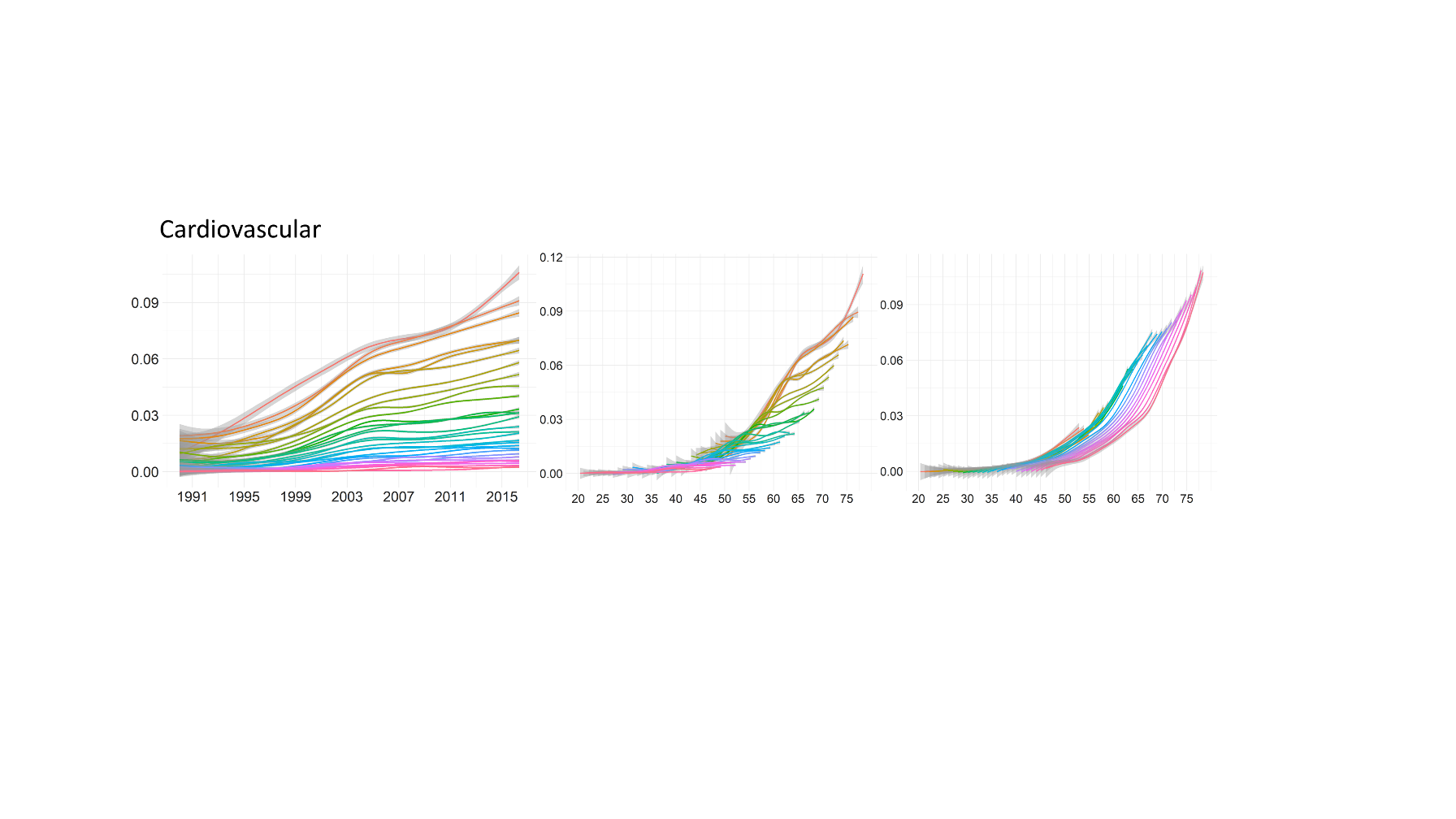

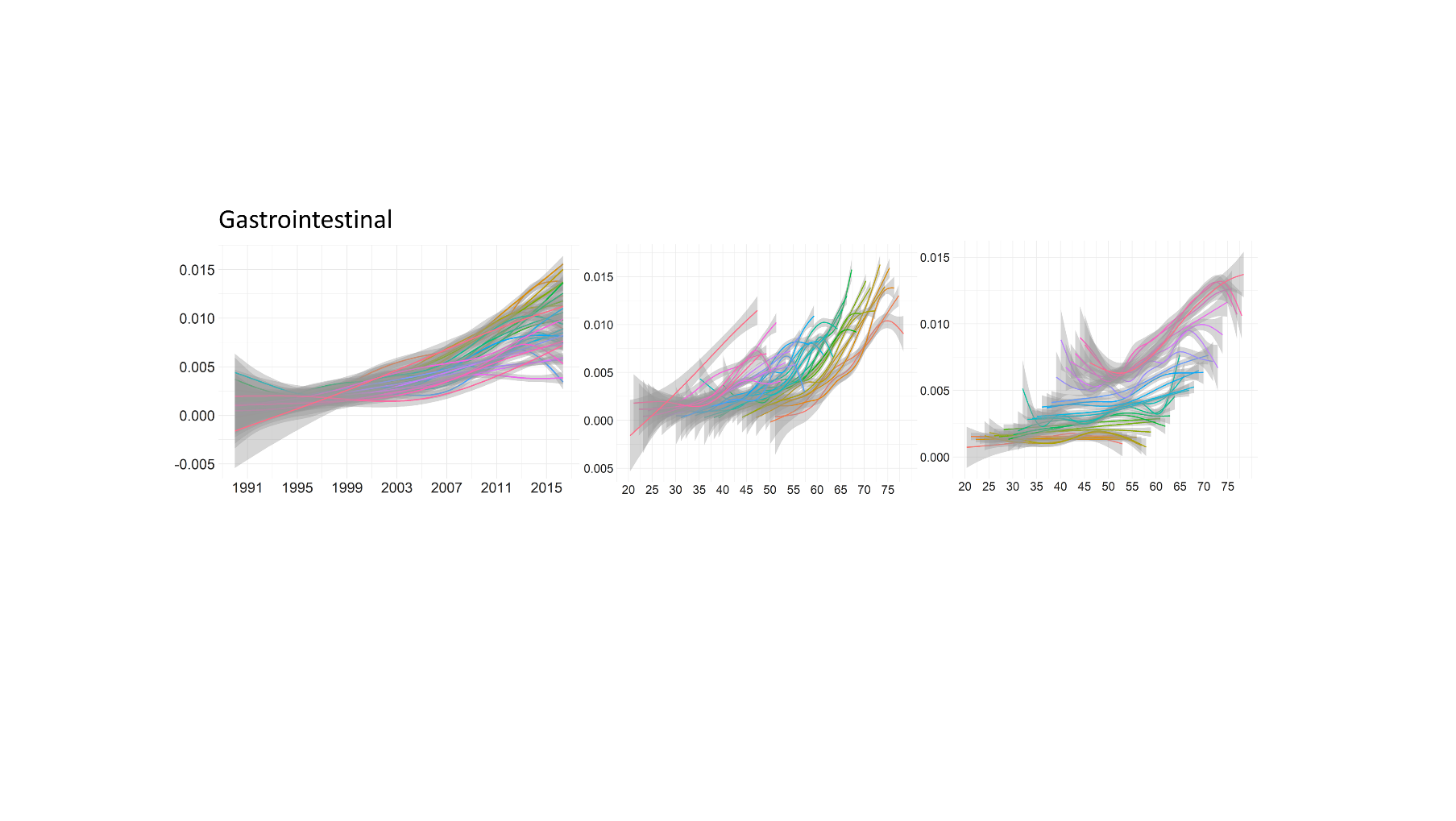

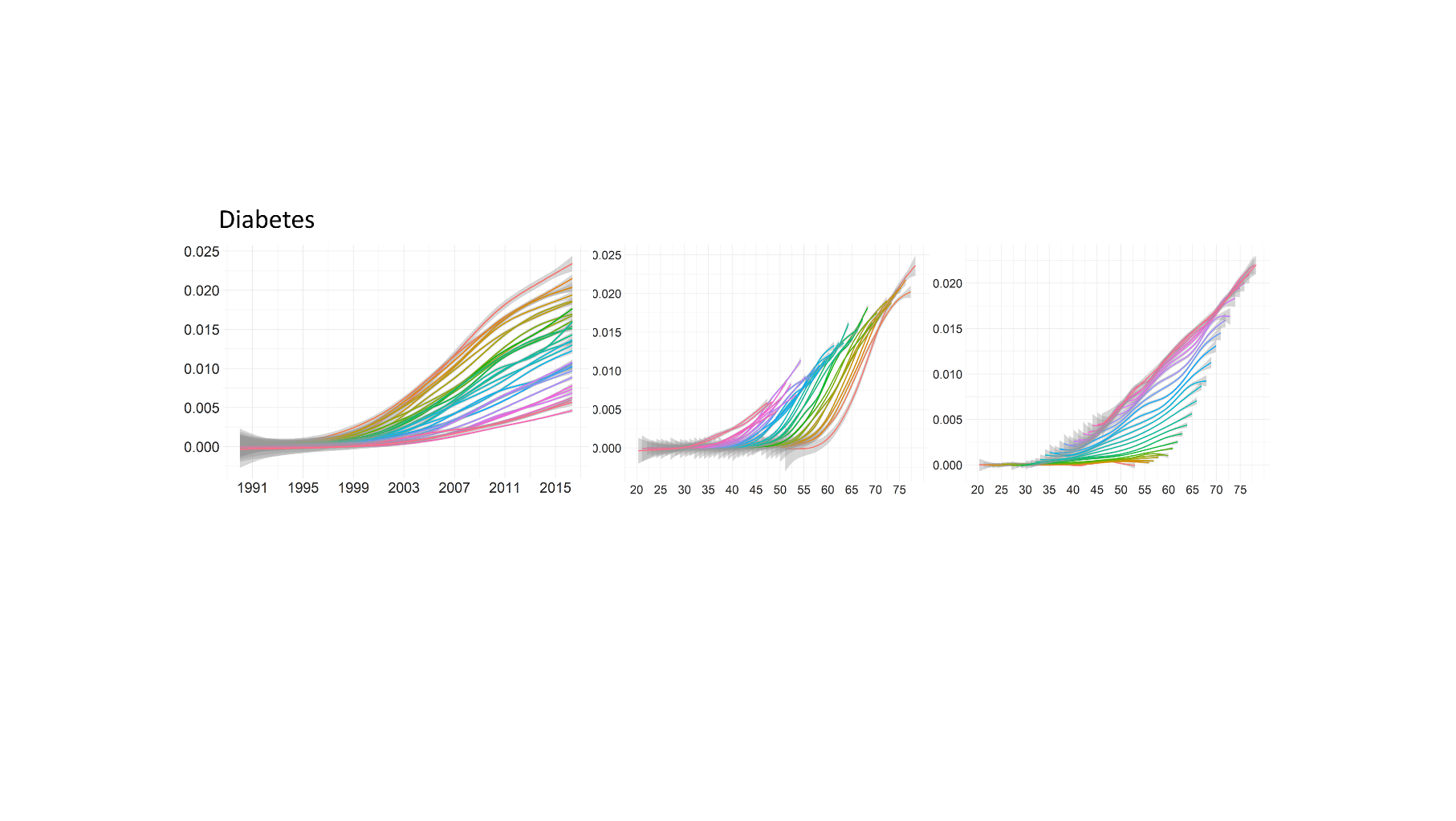

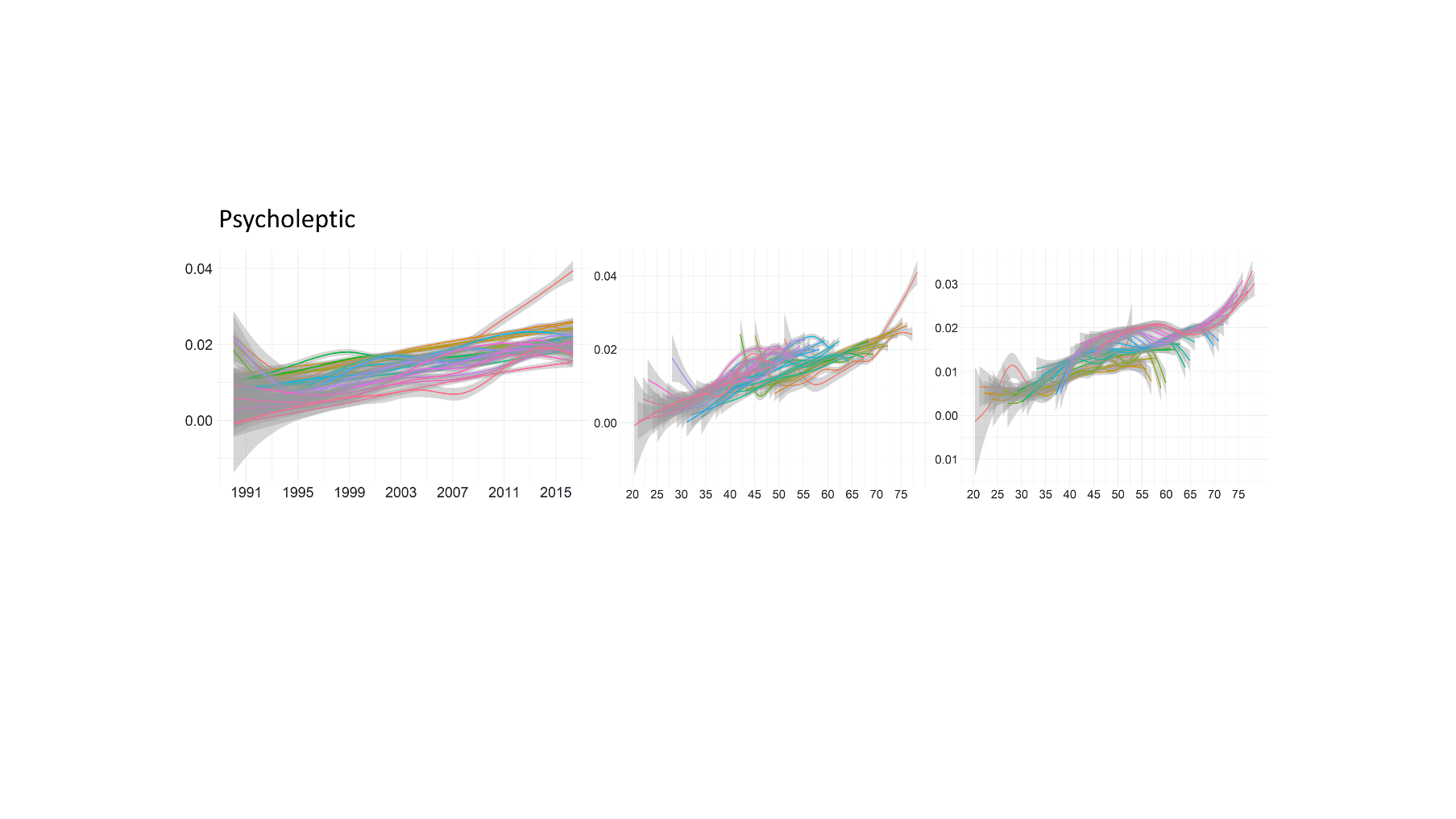

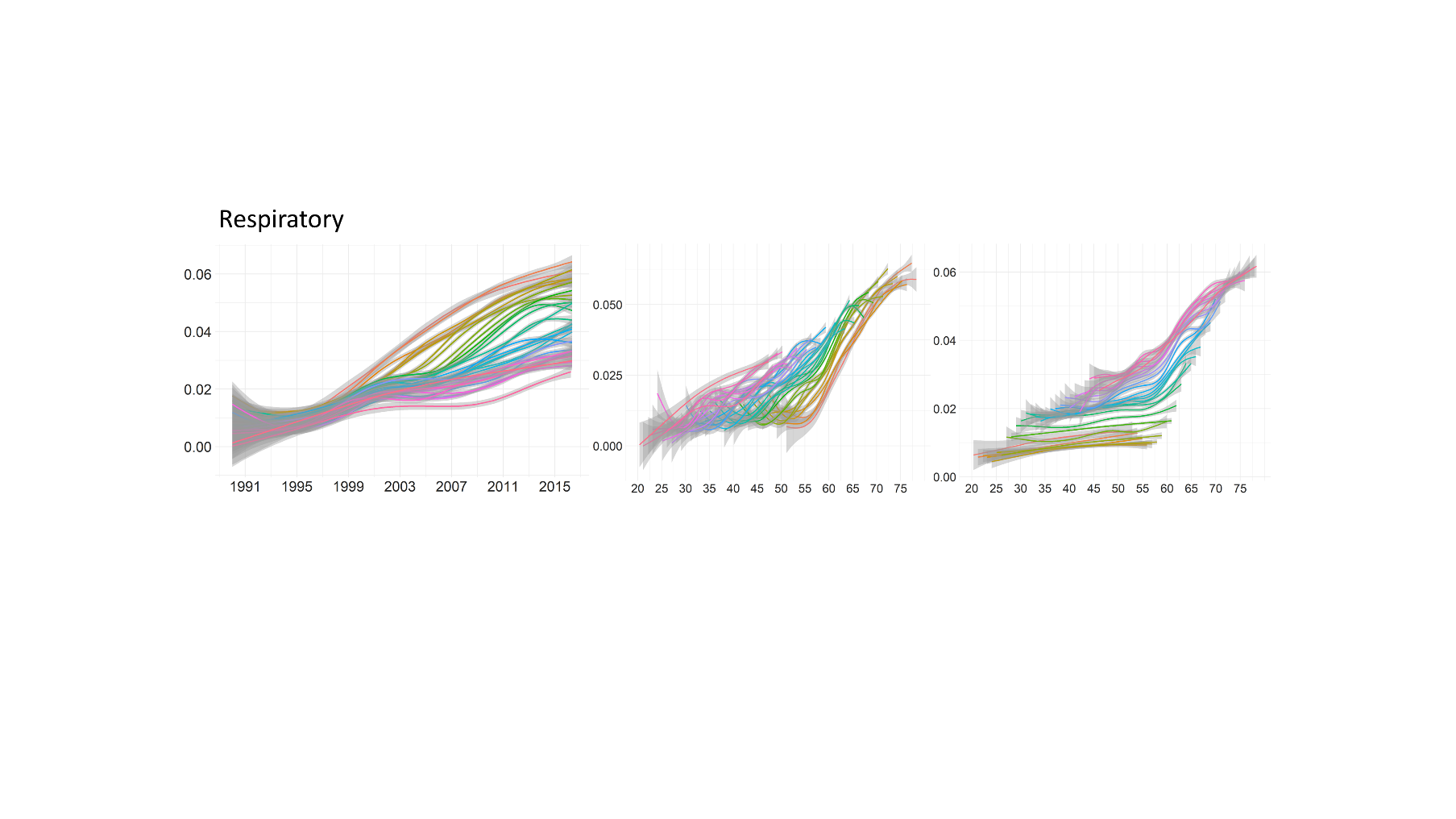

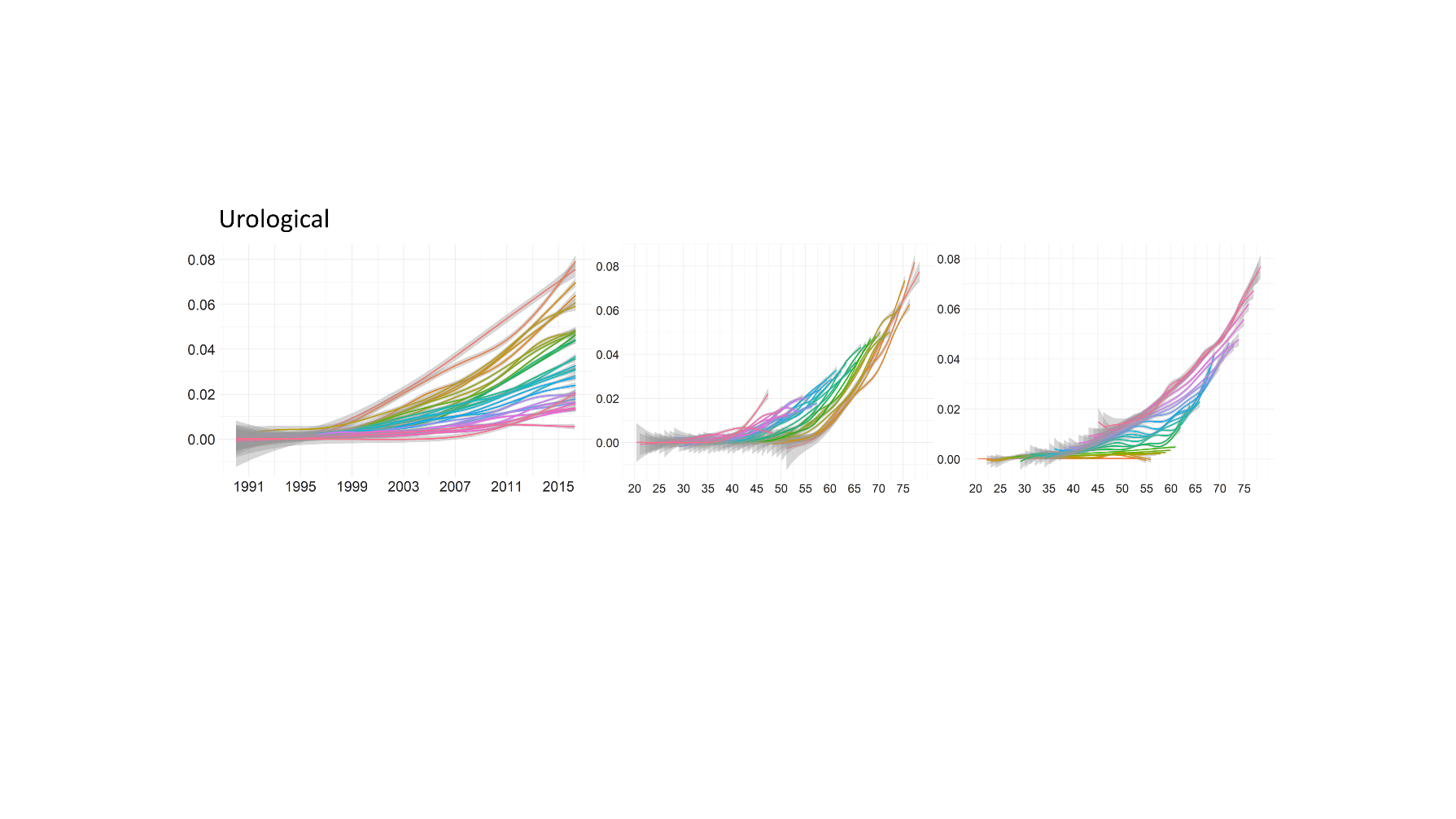

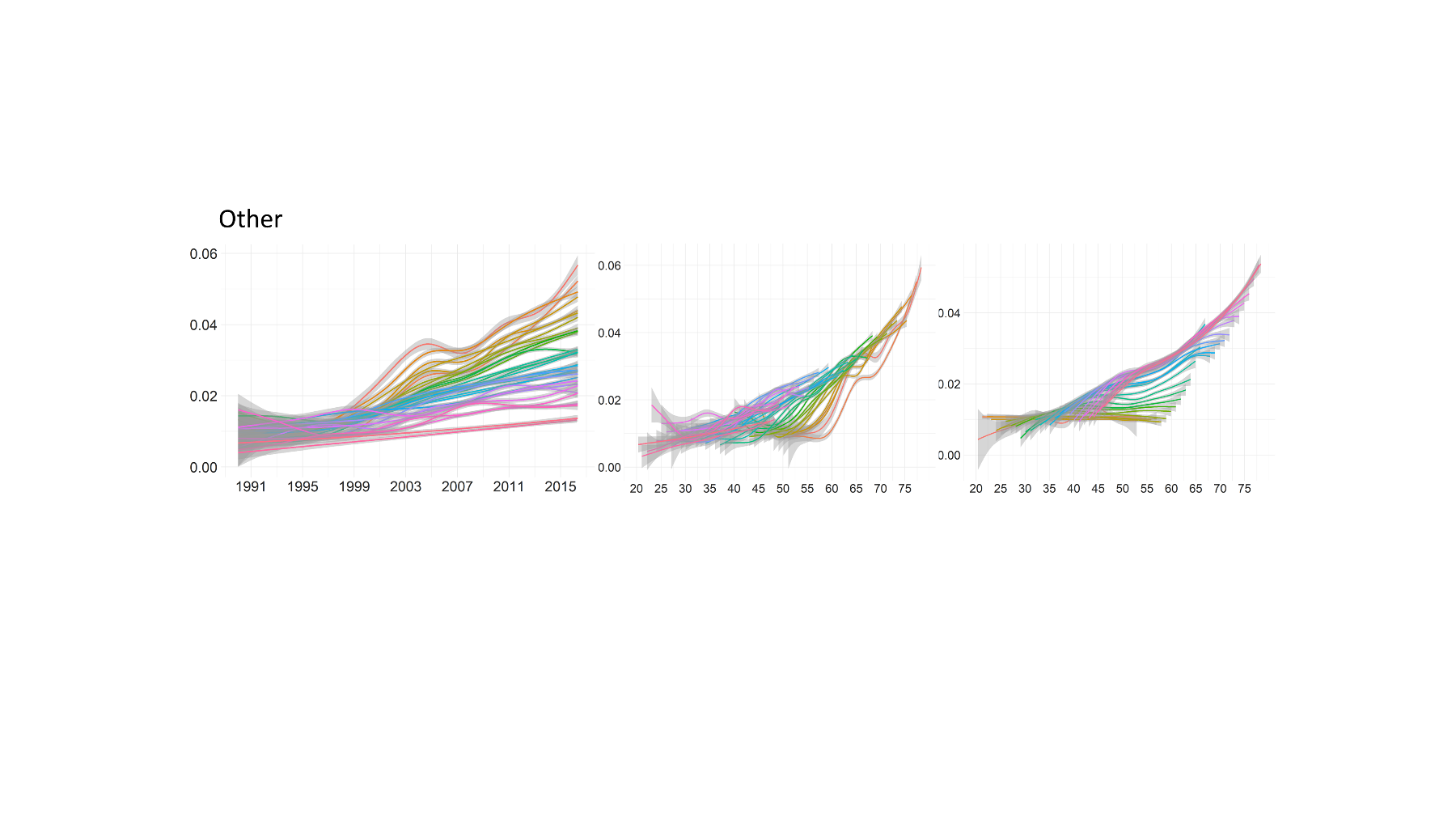

**Figure S7**: Plot of the number of prescribed anticholinergic drugs as a proportion of all drugs for each age group across the entire prescribing period. The x-axis represents the rounded age at time of prescription, the y-axis represents the ratio of the number of prescribed anticholinergic drugs and the number of all prescribed drugs.

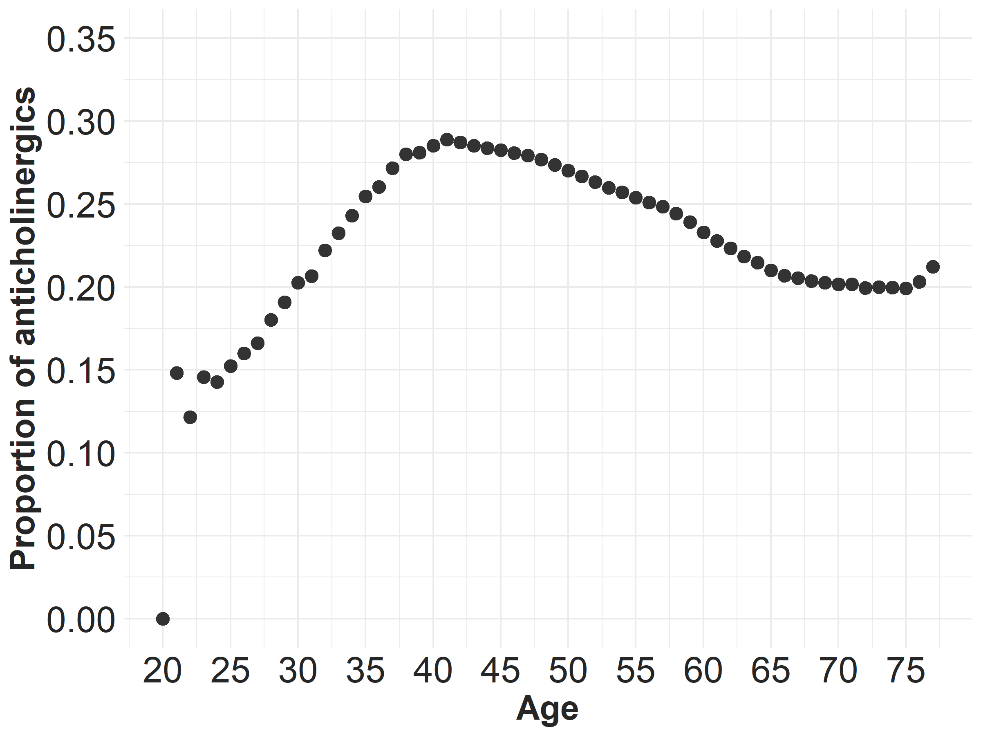

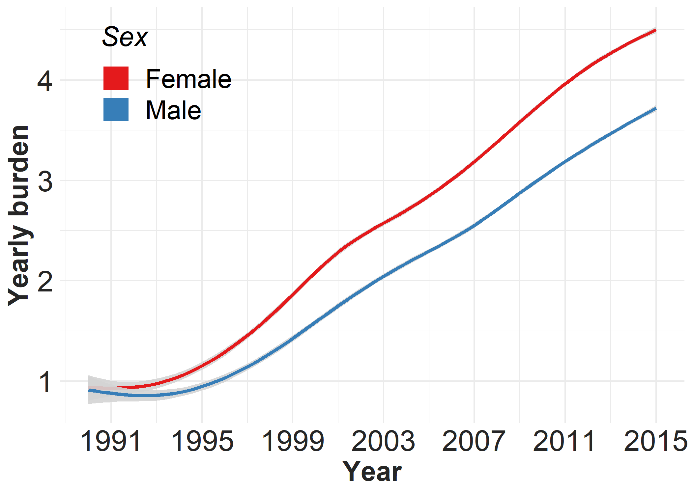

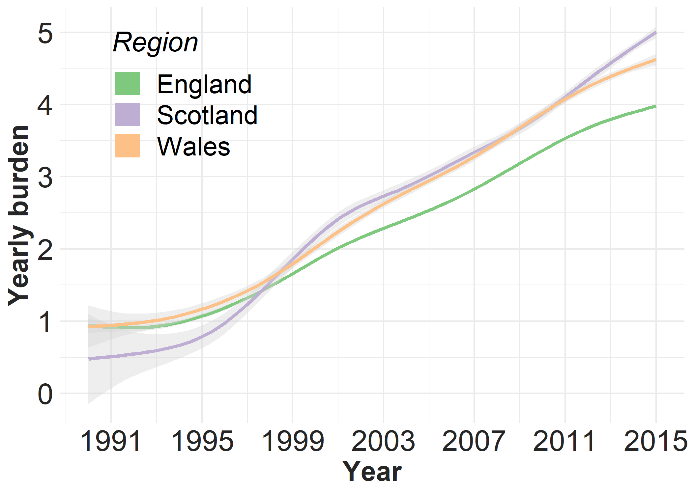

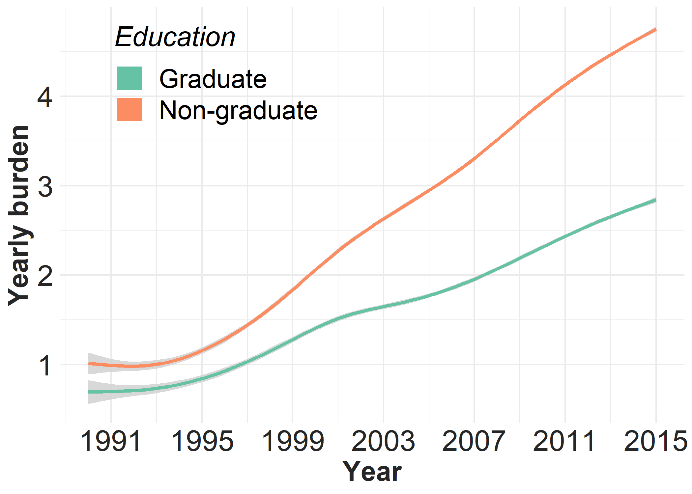

**Figure S8**: Temporal changes in anticholinergic burden for different levels of predictor variables. The plots were generated using generalised additive model smoothing. Shading indicates 95% confidence intervals.

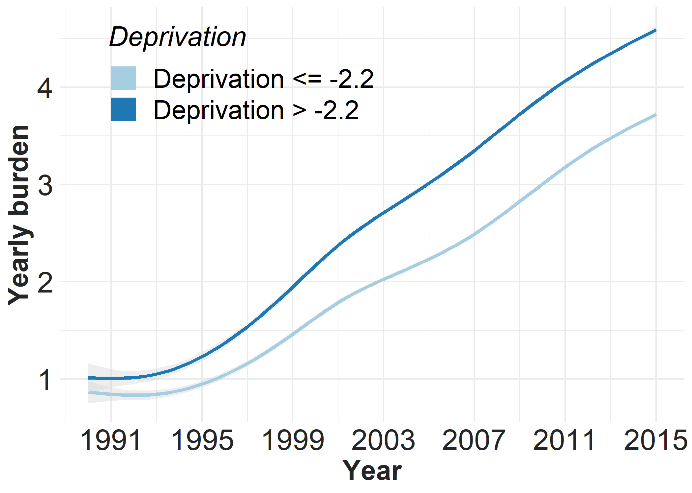
